# Brain atrophy progression in Parkinson’s disease is shaped by connectivity and local vulnerability

**DOI:** 10.1101/2021.06.08.21258321

**Authors:** Christina Tremblay, Shady Rahayel, Andrew Vo, Filip Morys, Golia Shafiei, Ross D. Markello, Ziv Gan-Or, Bratislav Misic, Alain Dagher

## Abstract

Atrophy in multiple brain regions has been reported in the early stages of Parkinson’s Disease, but there have been few longitudinal studies. How intrinsic properties of the brain, such as anatomical connectivity, local cell type distribution and gene expression combine to determine the pattern of disease progression remains unknown. One hypothesis proposes that the disease stems from prion-like propagation of misfolded alpha-synuclein via the connectome that might cause varying degrees of tissue damage based on local properties.

Here we used MRI data from the Parkinson Progression Markers Initiative to test this model by mapping the progression of brain atrophy over one, two and four years and relating it to brain structural and functional connectivity, cell type expression and gene ontology enrichment analyses. In this longitudinal study, we derived atrophy progression maps for the three time points using deformation-based morphometry applied to T1-weighted MRI from 74 de novo Parkinson’s Disease patients (50 Men: 24 Women) and 157 healthy control participants (115 Men: 42 Women). After regressing out the expected age and sex effects associated with normal aging, we found that atrophy significantly progressed over two and four years in the caudate, nucleus accumbens, hippocampus, and the temporal, parietal, occipital and posterior cingulate cortex. This progression was shaped by both structural and functional brain connectivity. Also, the progression of atrophy was more pronounced in regions with a higher expression of genes related to synapses and was related to the prevalence of oligodendrocytes and endothelial cells.

In sum, we demonstrate that the progression of atrophy in Parkinson’s Disease is in line with the prion-like propagation hypothesis of alpha-synuclein and provide evidence that synapses may be especially vulnerable to synucleinopathy. In addition to identifying vulnerable brain regions, this study reveals different factors that may be implicated in the neurotoxic mechanisms leading to progression in Parkinson’s Disease.

## Introduction

Atrophy has been reported in multiple brain regions even in the early stages of Parkinson’s disease.^1–3^ However, less is known about the progression of tissue loss after diagnosis.^4^ There have been a few studies of MRI-derived measures over time, but they were limited by small sample sizes,^5,6^ short follow-up durations,^7,8^ or use of global volumetric measures.^9–11^ The most consistent findings are subcortical tissue loss early on, notably affecting striatum, thalamus, amygdala and hippocampus, and more widespread cortical atrophy when longer durations or larger sample sizes are used. With few exceptions,^7^ no study has attempted to relate the spatial pattern of atrophy progression to intrinsic properties of the brain such as anatomical connectivity, cellular composition or regional gene expression. Such an analysis could provide information about pathophysiological mechanisms in Parkinson’s disease.

There is increasing evidence that brain connectivity may shape tissue loss in a multitude of neurodegenerative and psychiatric diseases including schizophrenia, frontotemporal dementia, amyotrophic lateral sclerosis, Alzheimer’s and Parkinson’s disease.^7,12–18^ This pattern points to the possibility of pathogenic agents spreading from neuron to neuron as a common mechanism. In Parkinson’s disease, converging evidence points to misfolded α-synuclein as the propagating agent.^19–24^ Most α-synuclein aggregates are localized in synapses where they are thought to interfere with neurotransmitter release and cause dendritic spine loss^25,26^ that may be detectable using MRI-based measures such as cortical thickness or deformation-based morphometry (DBM). Indeed, using a single time point or follow-up, we previously demonstrated that the MRI-derived baseline atrophy pattern and the atrophy progression after one year in *de novo* Parkinson’s disease are consistent with a connectome-based propagation,^2,7,16^ a finding that has been replicated computationally in animal models.^24^ However, the role of connectivity in explaining the longitudinal progression of atrophy in Parkinson’s disease has not yet been tested in human patients followed longitudinally with multiple MRI time points.

The progression of brain atrophy is likely not only explained by brain connectivity but also by regional variations in tissue vulnerability.^16,27^ Indeed, some brain areas with higher α-synuclein concentration appear to be more susceptible to injury but also more likely to act as disease propagators.^16,28,29^ Regional vulnerability may additionally depend on the local cellular distribution or gene expression patterns that render the milieu to be more neurotoxic or neuroprotective.^30–32^ For instance, microglia have been implicated in both neuroprotection and neurotoxicity via regulation of inflammation.^32,33^ Astrocytes have been shown to accumulate α-synuclein and produce proinflammatory cytokines and chemokines.^31^ Oligodendrocytes may be targeted by neurodegeneration but can also play a role in neuroprotection via the synthesis of neurotrophic factors.^34^ Endothelial cells have also been implicated in the progression of neural damage in Parkinson’s disease^35^ and blood-brain barrier (BBB) dysfunction is thought to play a role in the pathology associated with Parkinson’s disease and other neurodegenerative diseases.^36,37^ On the other hand, endothelial cells have also been shown to contribute to neuron survival under certain physiological and inflammatory conditions.^38,39^ Finally, the ratio of excitatory to inhibitory neurons could be relevant for disease pathology, as excitotoxicity has also been implicated in Parkinson’s disease, possibly in synergy with glial dysfunction.^40^ A virtual histology approach may help explain how cellular composition relates to local vulnerability to atrophy.^41^

How brain architecture, local cell type distribution and gene expression combine to determine the pattern of disease progression remains unknown. Here, we first mapped the progression of brain atrophy using DBM and then related it to structural and functional networks, cell type composition, and gene ontology enrichment analyses. We hypothesized that the constraints imposed by structural and functional networks as well as the regional differences in cell type composition would jointly shape the pattern of atrophy found in Parkinson’s disease. We show that the brain’s connectome significantly shapes the course of atrophy over four years and that oligodendrocytes and endothelial cells may play a role in neuroprotection, while gene ontology analysis points to synapses as an important target of neurodegeneration.

## Materials and methods

### Data acquisition

All clinical and structural MRI data were downloaded in July 2019 from the Parkinson’s progression markers initiative (PPMI, http://www.ppmi-info.org), a longitudinal multi-center database including *de novo* Parkinson’s disease participants and healthy controls (HC).^42^ Clinical measures and T1-weighted MRI acquired on 3 and 1.5 Tesla scanners at baseline, one, two, and four years were used in this study (see Table 1). Acquisition parameters and detailed protocols are described on the PPMI website (http://www.ppmi-info.org/wp-content/uploads/2018/02/PPMI-AM-13-Protocol.pdf). For each analysis, participants with missing data at one or more time points were excluded. Each participating center received approval from a local research ethics committee. All the procedures and tests followed these committees’ guidelines. Informed consent was obtained from each participant according to the Declaration of Helsinki before the beginning of the study.

**Table 1.** Descriptive statistics for the 74 *de novo* Parkinson’s disease patients

| Characteristics | Baseline<br>mean (SD) | Year 1<br>mean (SD) | Year 2 | Year 4 | F-test<br>p-Value | Bl -1Y<br>p-adjusted* | Bl -2Y<br>p-adjusted* | Bl -4Y<br>p-adjusted* |
| --- | --- | --- | --- | --- | --- | --- | --- | --- |
| Gender (men/women) | 50/24 | 50/24 | 50/24 | 50/24 | NA | NA | NA | NA |
| Number of patients taking PD medication | 0 | 55 | 66 | 71 | NA | NA | NA | NA |
| Age (years) | 60.2 (9.4) | 61.2 (9.4) | 62.3 (9.5) | 64.3 (9.5) | NA | NA | NA | NA |
| Education (years) | 15.2 (2.6) | NA | NA | NA | NA | NA | NA | NA |
| Disease duration (days) | 221.9 (228.7) | 602.4 (231.6) | 967.3 (228.9) | 1699.5 (234.4) | NA | NA | NA | NA |
| Motor symptoms |  |  |  |  |  |  |  |  |
| Hoehn and Yahr stage | 1.46 (0.50) | 1.64 (0.51) | 1.73 (0.53) | 1.78 (0.50) | < 0.00005 | 0.01 | 0.001 | < 0.00005 |
| MDS-UPDRS-III (total tremor) | 3.62 (3.02) | 4.06 (2.87) | 4.45 (3.55) | 4.68 (4.04) | 0.02 | 0.81 | 0.12 | 0.09 |
| MDS-UPDRS-III (total rigidity) | 3.64 (2.67) | 4.43 (2.88) | 4.91 (2.86) | 5.43 (2.86) | < 0.00005 | 0.02 | < 0.00005 | < 0.00005 |
| MDS-UPDRS-III (total) | 19.14 (8.53) | 20.83 (10.08) | 22.09 (11.32) | 23.01 (9.40) | 0.002 | 0.57 | 0.07 | 0.004 |
| Non-motor symptoms |  |  |  |  |  |  |  |  |
| Epworth sleepiness scale score | 4.19 (2.80) | 4.16 (2.91) | 4.55 (3.27) | 4.67 (3.47) | 0.23 | NA | NA | NA |
| SCOPA-autonomic | 8.58 (5.01) | 10.13 (5.85) | 10.91 (5.46) | 11.94 (5.83) | < 0.00005 | 0.02 | < 0.00005 | < 0.00005 |
| Montreal Cognitive Assessment | 27.40 (2.30) | 27.04 (2.65) | 26.78 (2.56) | 27.28 (2.72) | 0.12 | NA | NA | NA |
| Phonemic Fluency (Letter F)** | 12.33 (4.15) | 13.97 (4.88) | 14.07 (4.93) | 14.87 (4.78) | < 0.00005 | 0.045 | 0.02 | < 0.00005 |
| Semantic Fluency Mean (fruit. animal. vegetables)** | 16.45 (3.50) | 16.36 (3.62) | 16.86 (4.13) | 16.98 (4.35) | 0.24 | NA | NA | NA |
| HVLT-Total Recall | 25.32 (5.36) | 24.71 (5.85) | 24.43 (5.86) | 24.07 (6.11) | 0.14 | NA | NA | NA |
| HVLT-Delayed Recall | 8.56 (2.62) | 8.69 (2.74) | 8.65 (3.10) | 8.28 (3.23) | 0.5 | NA | NA | NA |
| Letter-Number Sequencing | 11.09 (2.75) | 10.77 (2.55) | 10.72 (2.98) | 10.49 (3.43) | 0.26 | NA | NA | NA |
| Mood |  |  |  |  |  |  |  |  |
| GDS-15 (Depressive symptoms) | 1.99 (2.17) | 2.07 (2.10) | 2.32 (2.80) | 2.26 (2.18) | 0.64 | NA | NA | NA |
| STAI (Anxiety) | 93.59 (8.25) | 91.96 (7.52) | 91.68 (7.61) | 92.99 (8.01) | 0.22 | NA | NA | NA |
| CSF Biomarkers (pg/ml) |  |  |  |  |  |  |  |  |
| Alpha Synuclein (α-syn) | 1455 (703) | 1423 (666) | 1412 (650) | NA | 0.84 | NA | NA | NA |
| Beta-Amyloid (Aβ) | 852 (354) | 865 (366) | 899 (354) | NA | 0.28 | NA | NA | NA |
| Phospho-tau (pTau) | 13.27 (4.84) | 13.24 (5.26) | 13.44 (4.73) | NA | 0.76 | NA | NA | NA |
| Total Tau (tTau) | 156 (52) | 158 (59) | 161 (51) | NA | 0.51 | NA | NA | NA |
| Neurofilament light polypeptide (NFL) | 11.3 (6.3) | 12.6 (7.3) | 13.6 (7.5) | NA | 0.001 | 0.002 | 0.001 | NA |

\*Bonferroni Correction
\*\*Number of words in one minute
Bl: Baseline; 1Y: One year from baseline; 2Y: Two years from baseline; 4Y: Four years from baseline
MDS-UPDRS: Movement Disorder Society Unified Parkinson Disease Rating Scale
SCOPA: SCAles for Outcomes in Parkinson’s disease
HVLT: Hopkins Verbal Learning Test
GDS: Geriatric Depression Scale; STAI: State-Trait Anxiety Inventory
CSF: Cerebrospinal Fluid; SD: Standard Deviation

### Participants

Table 1 describes the clinical characteristics of the participants with Parkinson’s disease included in the present study at each time point (i.e., baseline, one, two, and four years). Participants met the inclusion criteria for PPMI (http://www.ppmi-info.org/study-design/) described in the Supplementary Material (p.1). Participants with a history of Parkinson’s disease medication use, including L-Dopa, dopamine agonists, monoamine oxidase B inhibitors, or amantadine, within 60 days of the baseline visit, or with a diagnosis of dementia^43^ were excluded from the study. HC participants in the PPMI database were aged ≥30 years old at the screening visit with no current neurological disorder. Exclusion criteria for HC were a Montreal Cognitive Assessment (MoCA) score ≤26 or a first-degree relative with idiopathic Parkinson’s disease. There was a total of 403 patients with Parkinson’s disease and 184 HC with a T1-weighted MRI at baseline. After visual inspection to exclude images with abnormalities, 74 patients (50 men, 24 women) had both MRI and clinical evaluation at each of the three follow-up time points and 157 HC (115 men, 42 women) at baseline. Most exclusions were due to missing assessments at one of the time points. In addition, neuroimaging data passing quality control were available for 120 Parkinson’s disease participants at baseline and one year and 109 at two years, and these were used to confirm our findings with larger sample sizes. We ensured that the control group was similar to the Parkinson’s disease group (*n*=74) relative to age at baseline (HC mean = 60.1 y, range: 31-83; Parkinson’s disease patients mean = 60.2 y, range: 38-82 y; *p*=0.96) and sex (*χ^2^* = 0.09, *p* = 0.77).

We also assessed the sample for attrition bias, whereby more severely affected individuals tend to drop out of a longitudinal study. We used our previous classification of the patients in this dataset into three severity groups based on their baseline brain atrophy, which yielded mild, intermediate, and severe subtypes.^44^ The drop-out rates were 38%, 39%, and 58% for mild, intermediate and severe subtypes at two years (*χ^2^*=5.8, *p*=0.05) and 60%, 57%, and 70% at four years (*χ^2^*=2.5, *p*=0.30). This suggests that our dataset may have included less severely affected participants (compared to the entire *de novo* sample), especially at the two-year time point. Such a bias could obscure associations (e.g. between symptom severity and brain atrophy).^45^

### Brain structural analysis

#### Deformation based morphometry

DBM quantifies voxel-wise brain tissue atrophy by performing non-linear transformations from the participant’s brain to a template brain. DBM maps were derived from each participant’s T1-weighted MRI image at each time point using the Computational Anatomy Toolbox (CAT12)^46^ implemented in Statistical Parametric Mapping software (SPM12) (see Supplementary Material for the detailed protocol, p.2). Every voxel value represents the factor by which each voxel of the participant’s brain has to expand (positive value) or shrink (negative value) to be registered to the MNI template.

W-score maps were generated to account for age and sex on brain deformation.^47,48^ A W-score map was computed for each time point (baseline, one, two, and four years) using the following formula, at each voxel:

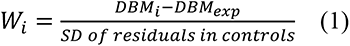

where *W_i_* is the W-score at voxel *i* for a participant, *DBM_i_* is the computed DBM value at voxel *i*, *DBM_exp_* is the expected DBM value for that participant defined by (*β1*\*age + *β2*\*sex + *β3*), and derived from HC data. All statistical analyses involving DBM were performed using the W-scored DBM maps. Negative W-score values indicate atrophy (reduced volume) whereas positive W-scores indicate expansion, with normal aging and sex effects accounted for. Only the baseline HC data were used as a reference since only a limited number of HC had longitudinal measurements.^49^

#### Brain atrophy progression analysis

The pattern of atrophy progression was investigated using two SPM12 toolboxes: CAT12^46^ and the probabilistic approach for threshold-free cluster enhancement (pTFCE)^50^, written in MATLAB (R2018b). A two-tailed repeated measures ANOVA (F-contrast) with time as covariate followed by post-hoc paired comparisons (t-contrasts between baseline (Bl) - 1 year, Bl - 2 year, Bl - 4 year) combined with the pTFCE based on Bayes’ rule were used to compare atrophy differences between time points in the Parkinson’s disease group. In all cases, W-score maps were used. Although sex was accounted for in the W-score calculation, we added it as a covariate in the repeated measures ANOVA to regress out its effect, as sex differences in atrophy were recently demonstrated in Parkinson’s Disease.^49^ Scanner site was also added as a covariate. Family-wise error (FWE) rate (threshold: *p_FWE_* < 0.05) was used to correct for multiple comparisons. Only significant clusters with 10 voxels or more were retained. This analysis was computed for the 74 participants with a T1-MRI at each of the four time points. Additional analyses (t-contrasts between Bl - 1 year or Bl - 2 years) were done with all the patients who had a T1-MRI at either baseline and one year (*N*=120) or baseline and two years (*N*=109).

To understand how tissue changes relate to brain function, the mean atrophy progression was computed for each of the seven cortical resting-state networks defined by Yeo et *al.*.^51^ These networks are thought to represent the distributed neural systems that support diverse cognitive domains.^52^ One-sample permutation tests were computed to compare the mean atrophy progression of each network to its null distribution (two-sided p-value). Statistical significance was estimated by permutation testing using the netneurotools toolbox (https://netneurotools.readthedocs.io/en/latest/index.html): network labels were permuted 1000 times while preserving the spatial autocorrelation, and network-specific means were recalculated to generate a null distribution for each network.^53,54^

To investigate the relationship between atrophy progression and the synaptic organization and hierarchy of different zones of the cortex, the cortical regions were defined following the nomenclature of Mesulam.^55^ The mean atrophy progression scores for each cortical tissue type (paralimbic, heteromodal, unimodal and idiotypic) were compared using a one-way ANOVA and post-hoc tests with Bonferroni corrections.

Correlations were computed to investigate the relationship between clinical measures (motor and non-motor symptoms, mood and CSF biomarkers) and atrophy progression in the whole brain and in each of the seven resting-state networks (threshold: *p_FDR_*<0.05, two-tailed). Partial Spearman’s correlations were used due to some data being non-normally distributed, with age, sex, and education as covariates (only age and sex were used for motor symptoms).

### Structural and functional network analysis

We next tested investigated the network spread hypothesis of Parkinson’s disease. We tested whether the atrophy progression observed in each region was correlated with the atrophy progression of its structurally and functionally connected neighborhood. In brief, 1) the brain was parcellated into equally sized cortical regions at four different spatial resolutions,^56^ 2) the mean atrophy progression was calculated for each region, 3) matrices of structural and functional connectivity between regions were constructed, using diffusion-weighted MRI tractography and resting state functional MRI acquired on a different dataset of 70 healthy adults,^57^ 4) the collective neighborhood atrophy progression of each region was calculated, using the structural and functional networks to define the neighbors, 5) correlations were computed between the atrophy progression in each region and its collective neighborhood atrophy progression, and 6) the significance of the correlations was tested against a null model preserving spatial autocorrelation. (See the detailed protocol in the Supplementary Material, p.3-5.) This network analysis approach was previously used in schizophrenia.^15^

#### Complementary analysis on the role of functional connectivity

Additional analyses were performed to verify whether FC, without considering the structural connections between regions, is related to atrophy progression. Pearson’s correlations were calculated between the regional atrophy progression and the collective deformation weighted by the FC of 1) the non-structurally connected direct neighbors (i.e. only region pairs not demonstrating structural connectivity, as defined above) and 2) all the regions irrespective of their structural connections. These correlations were compared against spatial autocorrelation null models (1000 spins, two-tailed). The *cocor* package was used to statistically compare the magnitude of the correlations and calculate a Zou’s confidence interval.^58^

### Cell type analysis

#### Virtual histology

We next investigated if the atrophy progression was associated with the prevalence of specific cell types in the cortex, notably astrocytes, endothelial cells, microglia, excitatory and inhibitory neurons, oligodendrocytes and oligodendrocyte precursors. A virtual histology approach was used to correlate the neuroimaging data with cell-specific gene expression across brain regions.^59,60^ Every class of cell type was associated with a gene list first derived by Seidlitz et *al.*,^41^ from five single-cell RNA sequencing studies of postmortem human cortical samples^61–65^ (for the detailed protocol see Hansen *et al.*).^66^ To generate cortical maps for each cell type, the spatial expression patterns of these gene lists were derived from postmortem brain data from six donors available in the Allen Human Brain Atlas (AHBA) genetics dataset.^67^ Cell type distribution was computed for each region of the Cammoun atlas^56^ at four different resolutions (68, 114, 219 and 448 cortical regions) using the abagen toolbox (https://github.com/rmarkello/abagen) and the recommendations laid out by Arnatkeviciute *et al.*.^68,69^ Pearson’s correlations were calculated between the atrophy progression of each region and the region’s average gene expression of each cell class. All the correlations were tested against null models preserving spatial autocorrelation (1000 spins; two-tailed).^54^

### Gene ontology enrichment analysis

A gene ontology (GO) enrichment analysis was performed to explore the biological processes related to atrophy progression over two and four years using brain regional gene expression. To do this, we extracted the average gene expression value for all genes available in the AHBA genetics dataset (i.e., 15,633)^67^ for each of the 448 cortical regions of the Cammoun atlas using the abagen toolbox and following previous recommendations.^68,69^ For investigating the functions of the genes associated with atrophy, we only selected the genes whose expression significantly correlated with atrophy progression after false discovery rate (FDR) correction and when compared to a null model preserving the spatial autocorrelation (1000 spins; two-tailed). This yielded lists of genes whose expression pattern was positively or negatively correlated with atrophy progression at two (positive correlation: *N* = 619 genes, negative correlation: 1058 genes) and four years (positive correlation: *N* = 900 genes, negative correlation: 1546 genes). We next investigated if the proportion of GO terms for the genes correlated to atrophy (i.e., the target gene list) significantly differed from the proportion of GO terms found with all genes extracted from the AHBA (i.e., background gene list). To ensure that the results were not due to the choice of a particular classification system, two publicly available gene ontology platforms were used to obtain GO terms: the Gene Ontology enRIchment anaLysis and visuaLizAtion tool (GOrilla)^70^ and the PANTHER Classification System.^71^ Of the 15,633 genes available in the AHBA genetics dataset, 13992 and 14657 genes were associated with a GO term in the GOrilla (GO Process) and PANTHER (GO biological process) platforms, respectively. Supported gene IDs are available from the GOrilla (http://cbl-gorilla.cs.technion.ac.il) and PANTHER (www.pantherdb.org) websites. For both platforms, a statistical overrepresentation analysis was conducted with Bonferroni correction to control for multiple comparisons. Whereas a hypergeometric model was implemented in GOrilla, the Fisher’s Exact test was used in PANTHER.

### Data availability

MRI data used in this article are available for download at www.ppmi-info.org/data. All other datasets and software used are available from the sources cited in the Methods. The DBM atrophy progression maps will be made available on request.

## Results

### Clinical measures and CSF biomarkers

For each feature and CSF biomarker presented in Table 1, two-tailed repeated measures ANOVA with post-hoc comparisons and Bonferroni-Holm correction^72^ were used to evaluate the progression after one, two, and four years. All measures of motor severity worsened over time, namely Hoehn and Yahr stage (*p*<0.00005), total UPDRS-III (*p*=0.002), tremor (*p*=0.02) and rigidity (*p*<0.00005). At each follow-up, the Hoehn and Yahr stage and total rigidity score were significantly different from baseline while the total UPDRS-III showed a significant difference only after four years (*p*-*adjusted*=0.004). Of the non-motor measures, only the SCOPA-autonomic and phonemic fluency scores (Letter F from MoCA) showed a significant difference with time (*p-adjusted*<0.00005). Both scores were significantly different from baseline at each follow-up. While the autonomic score worsened with time, an improvement was observed in phonemic fluency possibly reflecting a medication and/or learning effect (note that participants were not on anti-parkinsonian medications at baseline). Among the CSF biomarkers, only the neurofilament light polypeptide (NFL), a possible biomarker of neurodegeneration,^73,74^ presented a significant increase with time (*p*=0.0001), more specifically after one (*p-adjusted*=0.002) and two years (*p-adjusted*=0.001).

### Brain atrophy

The DBM W-Score maps were used to analyze voxel-wise atrophy progression after one, two and four years in the participants with Parkinson’s disease. This analysis showed an effect of time (F-contrast, two-tailed) in 24 clusters. The neuromorphometrics atlas (http://www.neuromorphometrics.com) was used to localize the significant clusters (Supplementary Table 1), which were widely distributed throughout the brain. Atrophy progression was mostly found in the caudate, nucleus accumbens, hippocampus, amygdala, and the temporal, parietal, occipital and cingulate cortex.

### Post hoc analysis

Post hoc tests (t-contrasts, one-tailed) were also computed between baseline and the three follow-up time points. Figure 1 (A) shows the regions with a significant atrophy progression after two and four years. No significant clusters presented significantly greater atrophy after one year. After two years, 11 clusters were significant (Supplementary Table 2), located in the bilateral temporal lobe, left precuneus, right caudate, and angular gyrus. After four years, significant atrophy progression was found in 13 clusters (Supplementary Table 2), located in the right caudate, bilateral nucleus accumbens, and temporal, parietal, occipital and cingulate cortex. Small clusters in the superior and inferior frontal gyrus were also found.

**Figure 1.**
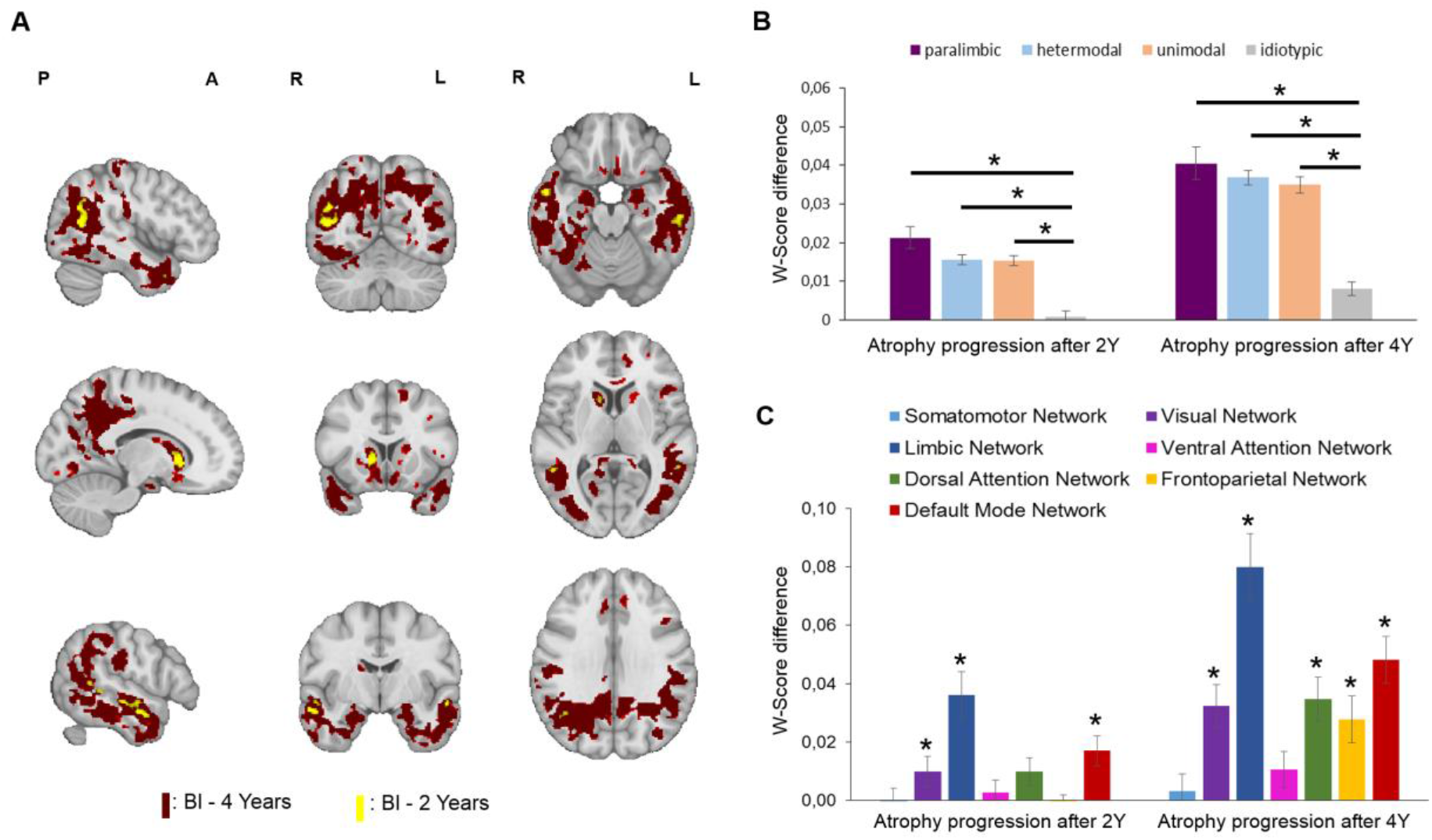
Atrophy progression after two and four years in *de novo* Parkinson’s disease patients. (**A**) Regions showing significant (*P_FWE_-value* < 0.05; one-tailed) differences between DBM W-Score at baseline (Bl), two (in yellow) and four years (in red) using the MNI152-2009c template for visualization purpose. (Top row: *x* = 45, *y* = -62, *z* = -19; Middle row: *x* = 14, *y* = 15, *z* = 7; Bottom row: *x* = 54, *y* = -6, *z* = 30) (B) Cortical regions were defined following the nomenclature of Mesulam based on synaptic organization and hierarchy. ^55^ The mean atrophy progression score and the standard error of the mean was calculated for each cortical zone. The idiotypic areas (primary sensory/motor cortex) showed a lower atrophy progression than the other cortical areas (\**p-value* < 0.05). (**C**) The atrophy progression pattern after two and four years was assigned to the seven resting-state networks defined by Yeo *et al.*.^51^ The mean atrophy progression score and the standard error of the mean was calculated for each network and compared to a null distribution (1000 spins; two-tailed) preserving spatial autocorrelation (\**p-value* < 0.05).

The previous results were derived from the participants who had scanning sessions at all time points. We repeated the 1-year and 2-year analyses using the larger sample sizes of individuals who have not undergone scanning at all time points. A high overlap was observed between the regions with a significant atrophy progression at two years in the full 4-year sample (described above) and the regions with atrophy progression found after including participants with a T1-MRI at baseline and one year (*N*=120), or baseline and two years (*N*=109) (Supplementary Fig. 1). Small clusters of atrophy progression were observed after one year in the additional analysis with 120 participants and larger clusters were found after two years with 109 participants (Supplementary Table 3).

To investigate the relationship with the synaptic hierarchy in the cortex, the average atrophy progression was compared between the paralimbic, heteromodal, unimodal and idiotypic cortical areas (Figure 1B) as defined by Mesulam.^55^ There was a significant effect of cortex type for the atrophy progression after two [*F*(3,995)=27.52, *p*<0.00005] and four years [*F*(3,995)=36.42, *p*<0.00005]. Post hoc comparisons indicated that this was due to significantly less atrophy progression in idiotypic (primary sensory and motor) cortex than other types. The mean atrophy progression values of the idiotypic areas after both two (*M*=0.0009, *SD*=0.02) and four year (*M*=0.008, *SD*=0.03) were significantly lower compared to the atrophy progression in other areas (*p-value*< 0.00005).

To further localize the disease process, the average atrophy progression was computed for each of the seven resting-state networks defined by Yeo *et al.*^51^ and compared against a null distribution preserving the spatial autocorrelation (1000 spins; two-tailed) (Figure 1C). The mean atrophy progression was significant after two years in the limbic (*p_spin_*=0.001), default mode (*p_spin_*=0.002), and visual (*p_spin_*=0.002) networks. After four years, the mean atrophy progression was also significant in the dorsal attention (*p_spin_*=0.001), frontoparietal (*p_spin_*=0.001), limbic (*p_spin_*=0.001), default mode (*p_spin_*=0.001) and visual (*p_spin_*=0.001) networks. Only the somatomotor (*p_spin_*=0.42) and ventral attention (*p_spin_*=0.10) networks did not present a significant atrophy progression after four years. Overall, the limbic and default mode networks were most affected.

### Relationship with clinical measures and CSF biomarkers

Correlation analyses were performed to investigate how the mean atrophy progression after two and four years related to 10 clinical characteristics (MDS-UPDRS-III (total), the non-motor features and depressive symptoms) and all the CSF biomarkers presented in Table 1. Partial Spearman’s correlations controlling for the effect of age, sex, and education were not significant before (*p*<0.01; two-tailed) or after FDR corrections (*p_FDR_*<0.05; two-tailed). When investigating the association between the mean atrophy progression of the seven resting-state networks after two and four years and the change in clinical measures over the same time period, no significant correlations were found after correcting for multiple comparisons.

### The influence of structural and functional connectivity

We next investigated if there was a network-specific distribution of atrophy progression. If this is the case, brain regions structurally and/or functionally connected with neighbors showing more atrophy progression should also present more atrophy progression after two or four years. For both structural and functional connectivity, significant correlations against spatial null models were found between the atrophy progression of a region and the atrophy progression of its connected neighbors after two years (structural: *r*=0.61, *p-value_spin_*=.0001; FC: *r*=0.62, *p-value_spin_*=0.0001; two-tailed) and four years (structural: *r*=.61, *p-value_spin_*=0.0001; FC: *r*=0.64, *p-value_spin_*=0.0001; two-tailed) at a parcellation of 448 cortical regions (Figure 2). The results were replicated at all other spatial resolutions (Supplementary Fig.2).

**Figure 2.**
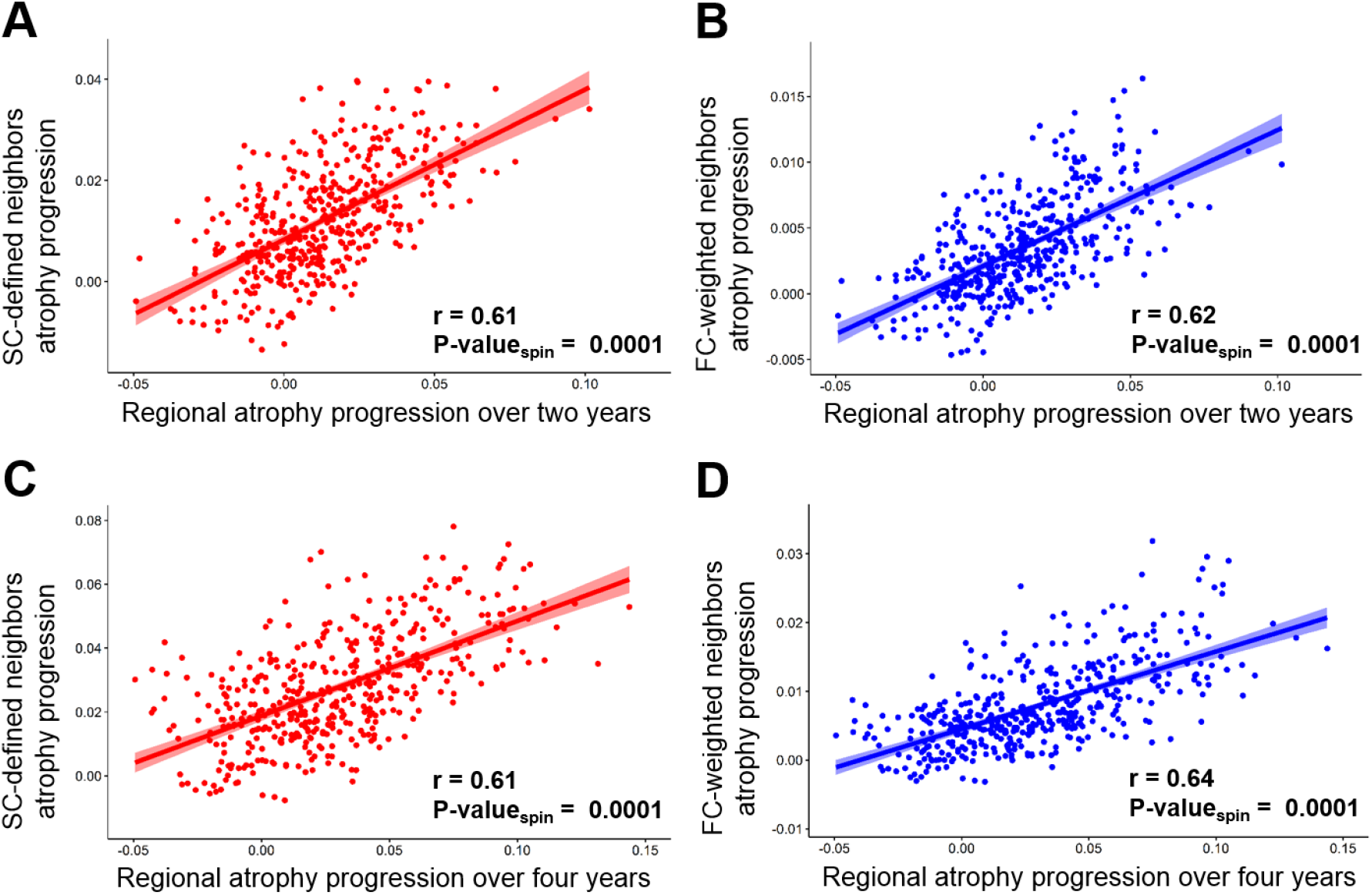
Relationship of atrophy progression to structural and functional connectivity. (**A, C**) Regional atrophy progression was significantly correlated with the atrophy progression of the structurally connected (SC) neighboring regions after two (**A**) and four years (**C**). (**B, D**) Regional atrophy progression was also related to the atrophy progression of the neighboring regions weighted by their FC after two (**B**) and four years (**D**). All the correlations showed were calculated using the Cammoun atlas with 448 regions, but similar results were obtained with three other resolutions (Supplementary Fig.2).

FC from resting state fMRI is constrained by structural connectivity.^75^ To verify more specifically its influence on atrophy progression, we investigated the relationships between the regional atrophy progression and the collective deformation weighted by the FC of 1) the non-structurally connected neighbors and 2) all the regions irrespective of their structural connections. Table 2 shows that, in both cases, correlations were markedly reduced. These results were replicated at the other spatial resolutions. Altogether, this finding suggests that both structural and functional connectivity are associated with atrophy progression, but that neuronal activity modulates co-atrophy between structurally connected regions.

**Table 2.**
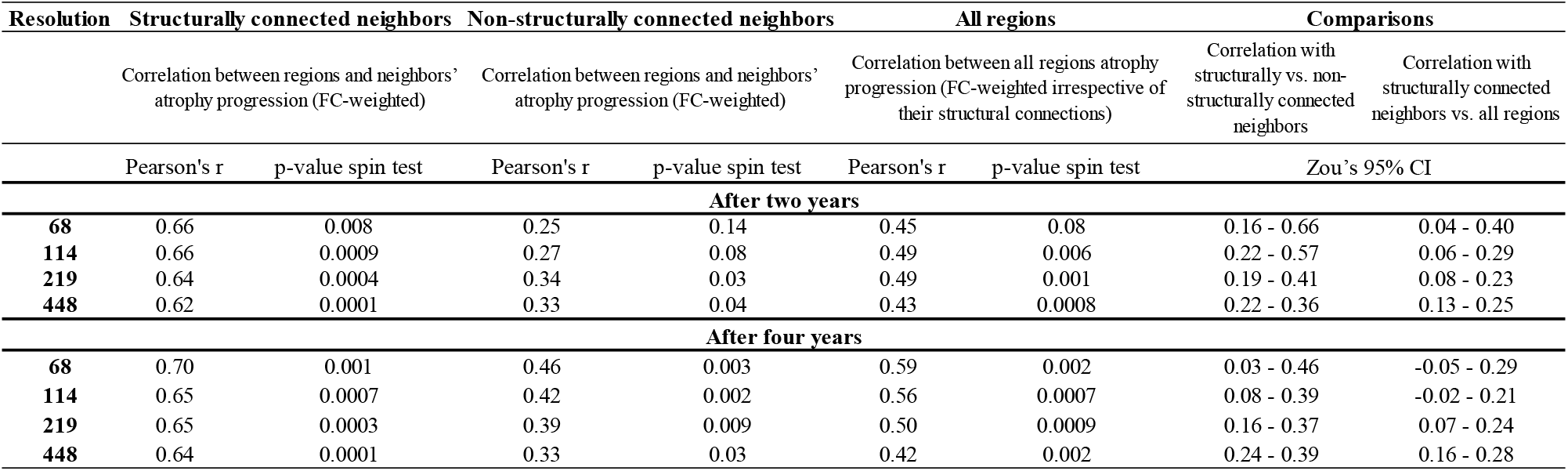
Influence of functional connectivity on atrophy progression

| Resolution | Structurally connected neighbors |  | Non-structurally connected neighbors |  | All regions |  | Comparisons |  |
| --- | --- | --- | --- | --- | --- | --- | --- | --- |
|  | Correlation between regions and neighbors' atrophy progression (FC-weighted) |  | Correlation between regions and neighbors' atrophy progression (FC-weighted) |  | Correlation between all regions atrophy progression (FC-weighted irrespective of their structural connections) |  | Correlation with structurally vs. non-structurally connected neighbors | Correlation with structurally connected neighbors vs. all regions |
|  | Pearson's r | p-value spin test | Pearson's r | p-value spin test | Pearson's r | p-value spin test | Zou's 95% CI |  |
|  | After two years |  |  |  |  |  |  |  |
| 68 | 0.66 | 0.008 | 0.25 | 0.14 | 0.45 | 0.08 | 0.16 - 0.66 | 0.04 - 0.40 |
| 114 | 0.66 | 0.0009 | 0.27 | 0.08 | 0.49 | 0.006 | 0.22 - 0.57 | 0.06 - 0.29 |
| 219 | 0.64 | 0.0004 | 0.34 | 0.03 | 0.49 | 0.001 | 0.19 - 0.41 | 0.08 - 0.23 |
| 448 | 0.62 | 0.0001 | 0.33 | 0.04 | 0.43 | 0.0008 | 0.22 - 0.36 | 0.13 - 0.25 |
|  | After four years |  |  |  |  |  |  |  |
| 68 | 0.70 | 0.001 | 0.46 | 0.003 | 0.59 | 0.002 | 0.03 - 0.46 | -0.05 - 0.29 |
| 114 | 0.65 | 0.0007 | 0.42 | 0.002 | 0.56 | 0.0007 | 0.08 - 0.39 | -0.02 - 0.21 |
| 219 | 0.65 | 0.0003 | 0.39 | 0.009 | 0.50 | 0.0009 | 0.16 - 0.37 | 0.07 - 0.24 |
| 448 | 0.64 | 0.0001 | 0.33 | 0.03 | 0.42 | 0.002 | 0.24 - 0.39 | 0.16 - 0.28 |

### Relationship with cell-type distribution

In addition to connectivity, local factors may also influence atrophy progression in Parkinson’s disease. Here we investigated the relationship between the relative regional prevalence of different cell types and cortical atrophy progression. Two cell types were associated with relatively less atrophy progression. Using the Cammoun atlas with 448 regions, significant negative correlations were found between the prevalence of endothelial cells and atrophy progression after two (*r*=-0.15, *p-value_spin_*=0.04, two-tailed) and four years (*r*=-0.19, *p-value_spin_* =0.01, two-tailed) (Figure 3). The atrophy progression was also inversely correlated with the prevalence of oligodendrocytes after two (*r*=-.11, *p-value_spin_*=0.049, two-tailed) and four years (*r*=-0.11, *p-value_spin_* =0.04, two-tailed). In addition, negative correlations were obtained with three lower resolutions (68, 114 and 219 regions (R)) (see Supplementary Fig.3). Only the correlations between the atrophy progression after four years and the endothelial cells were significant at the three other resolutions (68R: *r*=-0.33, *p-value_spin_* =0.04; 114R: *r*=-0.33, *p-value_spin_*=0.003; 219R: *r*=-0.25; *p-value_spin_*=0.049) while the correlations between the atrophy progression after four years and oligodendrocytes were near statistical significance (68R: *r*=-0.23; *p-value_spin_*=0.06; 219R: *r*=-0.14; *p-value_spin_*=0.06) or significant (114R: *r*=-0.21; *p-value_spin_*=0.049). No significant correlation was found for the other five cell types (astrocytes, microglia, excitatory and inhibitory neurons and oligodendrocyte precursors) (Supplementary Fig.4), and no cell type was associated with greater progression. These results seem to indicate slower cortical atrophy progression in regions with a higher prevalence of endothelial cells or oligodendrocytes.

**Figure 3.**
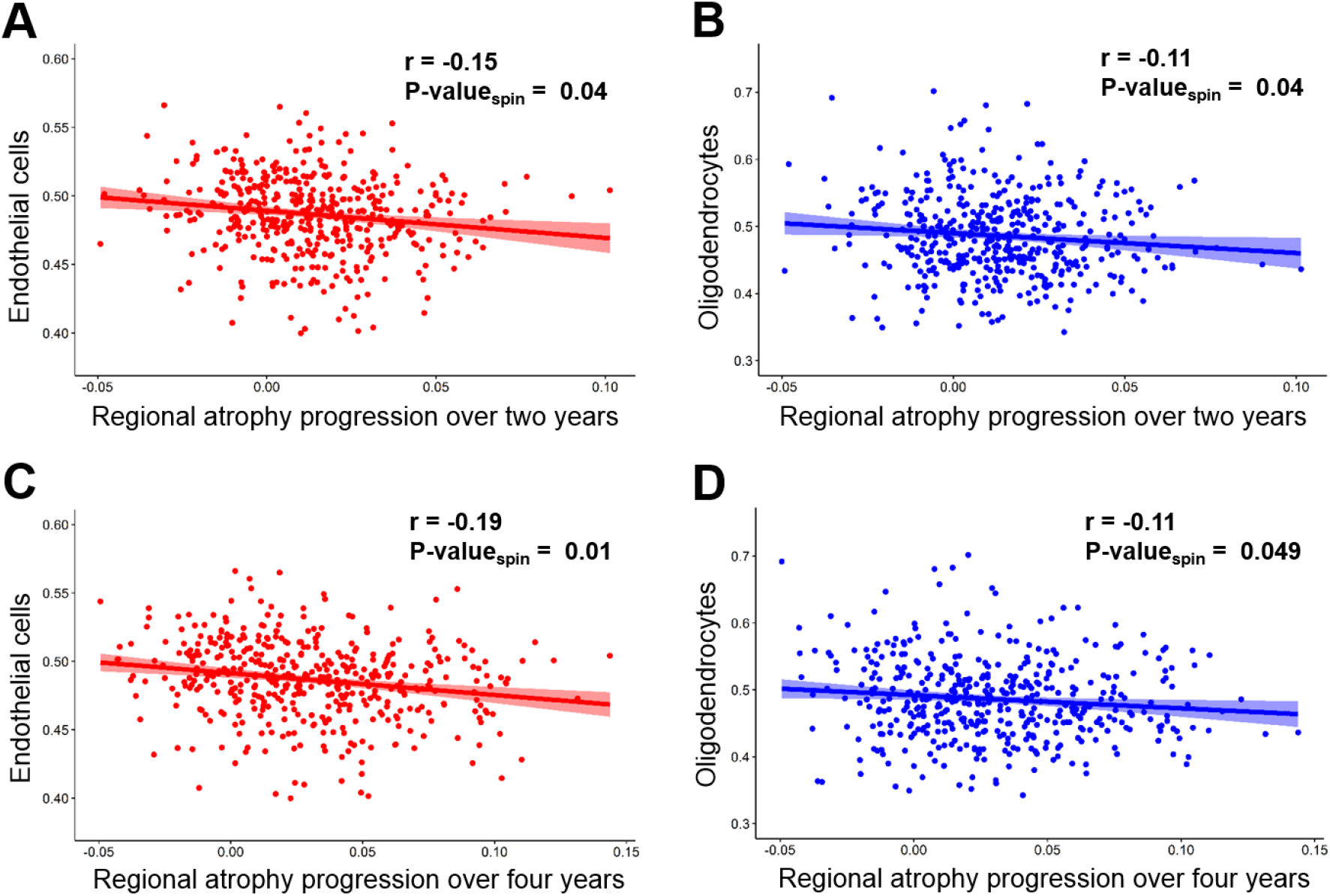
Specific cell-type prevalence related to atrophy progression in Parkinson’s disease. (**A, C**) The regional prevalence of endothelial cells in the brain was correlated with lower atrophy progression observed after two (**A**) and four years (**C**). (**B, D**) A negative correlation was also found with the prevalence of oligodendrocytes for atrophy progression after two (**B**) and four years (**D**). All the correlations showed were calculated using the Cammoun atlas with 448 regions, but negative correlations were also obtained with three lower resolutions (Supplementary Fig.3).

### Relationship with specific biological processes

To determine the functions of the genes whose expression was spatially associated with atrophy progression, a gene ontology (GO) enrichment analysis was done. Two platforms, Gorilla and PANTHER, were used. The results from each platform consistently implicated terms related to synaptic function. Figure 4 shows the significant GO terms (*p-value_Bonferonni_* <0.05, two-tailed) from the genes positively associated with the atrophy progression after two (*N*=23) and four years (*N*=17), and their average fold enrichment. Individual platform results are shown in Supplementary Fig.5 and Fig.6. The GO enrichment analysis revealed processes related to synaptic activity (regulation of synaptic plasticity Fig.4A-B, fold enrichment = 3.61 and 3.33), chemical synaptic transmission (Fig.4A-B, fold enrichment = 3.16 and 2.67) and cell signaling, namely trans-synaptic signaling (Fig.4A-B, fold enrichment = 3.12 and 2.62), and cell-cell signaling (Fig.4A-B, fold enrichment = 2.21 and 2.05) from the genes related to the atrophy progression after two (13 GO terms) and four years (10 GO terms). In sum, the GO analysis revealed that the regions showing greater atrophy progression tend to have greater expression of genes implicated in synaptic activity and cell signaling.

**Figure 4.**
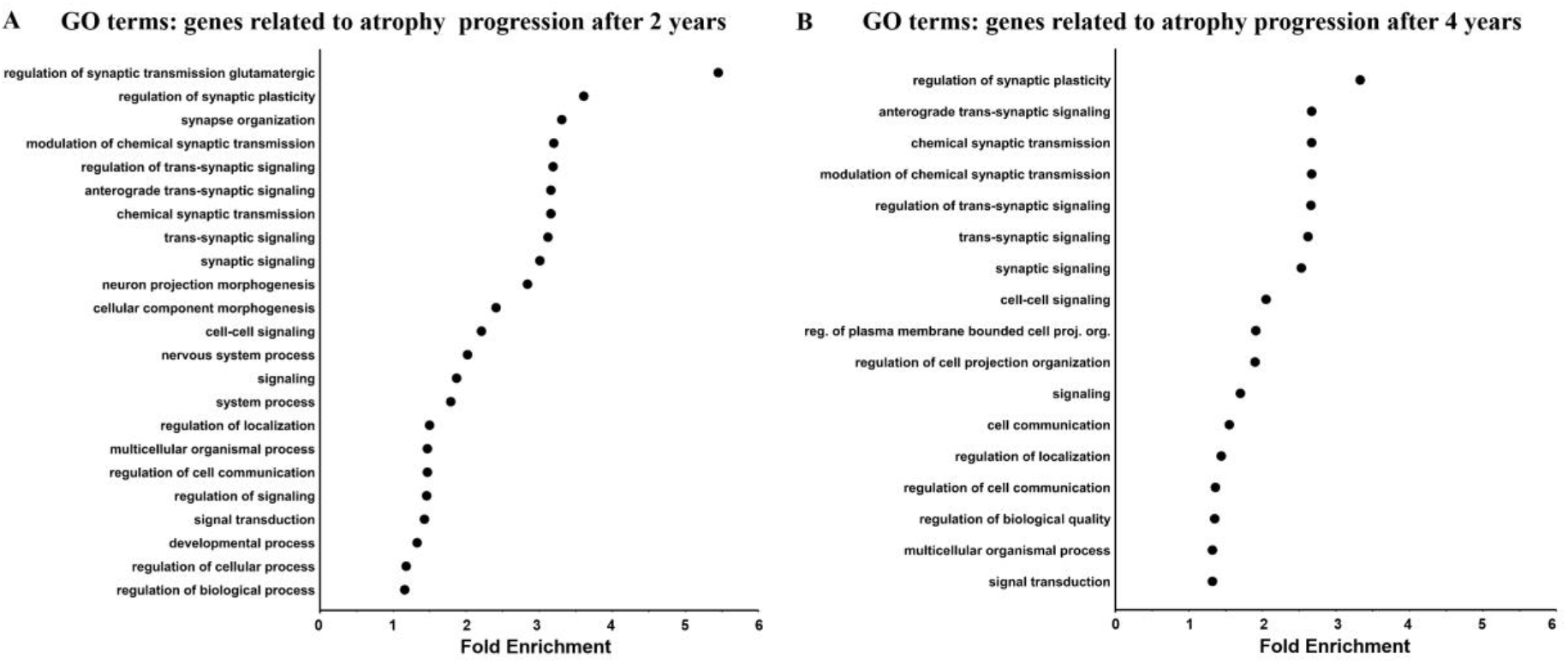
Gene ontology (GO) enrichment analysis of the genes related to atrophy progression in Parkinson’s disease. The fold enrichment was obtained by calculating the ratio of the genes positively related to atrophy progression after two (**A**) and four years (**B**) over the number of expected genes for each GO term, based on the background genes list. Only the significant results (*p-value_Bonferonni_* < 0.05) obtained with both platforms (GOrilla and PANTHER) are shown here. The average fold enrichment values are the average from the two platforms. There was no significant GO term for the genes negatively associated with atrophy progression at both time points with either platform.

Additional analysis shows that 33 genes uncovered in this analysis were located in known Parkinson’s disease GWAS loci^76^ (Supplementary Table 4 and Table 5), but future studies would be needed to examine whether these specific genes are driving the associations of the relevant loci with Parkinson’s disease atrophy.

## Discussion

Progressive brain atrophy was observed after two and four years in *de novo* Parkinson’s disease after regressing out the effects of sex and normal aging. The atrophy distribution was widespread, affecting five of the seven canonical resting-states networks^51^ at four years. In line with the Braak hypothesis,^19^ cortical atrophy progression was determined by brain connectivity, supporting trans-neuronal propagation of the pathogenic process. Atrophy was greater in regions enriched for genes related to synaptic activity and signaling. In addition, cortical regions containing relatively more oligodendrocytes and endothelial cells had reduced vulnerability to atrophy. These findings may point to the biological mechanisms at play in neurodegeneration in Parkinson’s disease. More generally, they support a model according to which neurodegeneration results from the interaction of a propagating agent with regional vulnerability.

### Brain atrophy progression

Evaluating the progression of atrophy in the whole brain using DBM measures, we found small regions over the temporal and parietal lobe in addition to the right caudate nucleus presenting an atrophy progression after two years, while a more widespread atrophy progression in the caudate nucleus, nucleus accumbens and the temporal, parietal, occipital and cingulate cortex was observed after four years. This mostly posterior cortical pattern of atrophy progression is similar to the atrophy distribution observed in the ENIGMA-Parkinson’s Study including 2,367 patients with Parkinson’s disease and 1,183 HC.^77^ Our findings showed alterations in the default mode, limbic, dorsal attention, frontoparietal and visual networks over this time. No relationship was found between the atrophy progression in any of the resting-state networks and the change in clinical symptoms. One explanation may be that the clinical measures available did not specifically target the regions showing atrophy progression. Indeed, motor deficits are mainly due to dopamine deficiency in the substantia nigra^78^ while autonomic dysfunction is mostly associated with brainstem and spinal neuronal loss in Parkinson’s disease.^79^ The substantia nigra may not show atrophy progression due to a floor effect (cell loss being already severe at presentation), while the brainstem is difficult to assess with DBM. Cortical atrophy patterns may be expected to relate to cognitive impairment, however, no cognitive decline was observed in this patient group, possibly due to a form of attrition bias affecting the PPMI cohort, whereby more severely affected individuals tended not to undergo the later MRI sessions. This may also explain why atrophy did not impinge on frontal areas in this sample. It is expected that more broadly distributed atrophy progression in later stages of the disease will eventually lead to cognitive deficits.^80^

### Structural and functional connectivity mediates atrophy progression

We have previously shown that atrophy in de novo Parkinson’s disease targets an intrinsic brain network^2^, supporting the network spread hypothesis. Here, atrophy progression significantly affected five of the seven resting-state networks; however, to test the role of connectivity we asked whether cortical regions with greater atrophy progression were more likely to be structurally and functionally connected with each other. Correlations were computed against spatial null models to control for the intrinsic relationships between proximal cortical regions. Our results indicate that both structural and functional connectivity are related to the atrophy progression after two and four years, even after controlling for regional autocorrelation in the brain. This is in line with studies suggesting that atrophy in Parkinson’s disease progresses via neuronal connectivity,^2,7,14,16,21^ most likely reflecting the spread of misfolded α-synuclein via neuronal projections.^19,22,23,81^ Moreover, our findings are in accordance with previous work showing that the atrophy pattern in other neurodegenerative follows structural and functional brain network architecture.^12,15,23,82^

Our previous work with an agent-based spreading model demonstrated that the pattern of brain atrophy depends on both neuronal connectivity and regional factors.^16^ Local vulnerability to neurodegeneration could depend on several factors including the prevalence of specific cell types ^30–32^. We next used gene-expression data to test for these local factors.

### Cellular composition may reflect regional vulnerability

Using gene expression associated with different cell types in the healthy brain,^41^ the relationships between the prevalence of seven cell types (astrocytes, endothelial cells, microglia, excitatory and inhibitory neurons, oligodendrocytes and oligodendrocyte precursors) and atrophy progression were investigated. Only significant negative correlations were observed between regional atrophy progression and the prevalence of oligodendrocytes and endothelial cells in the cortex, suggesting a possible neuroprotective effect of both of these cell types. Our finding is in line with studies showing that oligodendrocytes promote neuronal survival by secreting trophic factors such as brain-derived neurotrophic factor (BDNF), enhancing neuronal survival and axon regeneration, insulin-like growth factor 1 (IGF1), increasing the survival of young and aged cortical neurons, and glial cell line-derived neurotrophic factor (GDNF), known for its neuroprotective effect on dopaminergic neurons.^83–87^ However, little is known about the role of oligodendrocytes in Parkinson’s disease. A small number of α-synuclein inclusions has been observed in non-myelinating oligodendrocytes that may eventually contribute to neuronal death,^31^ but the production of neurotrophic factors by both myelinating and non-myelinating oligodendrocytes may override this neurotoxic effect. Oligodendrocyte genes have also been related to cortical synaptic density and likely play a role in synaptic elimination and maturation.^88^

The role of endothelial cells and the BBB in the progression of Parkinson’s disease is unclear. Some studies suggest that BBB alterations could contribute to neuropathological damage^36,37^ while others showed that endothelial cells contribute to neuronal survival under both physiological and inflammatory conditions.^38,39^ Endothelial cells are part of a neurovascular niche that supports neurogenesis in the adult brain.^89–91^ In addition, many angioneurins (including BDNF, IGF1 in addition to nerve, vascular endothelial, hepatocyte and epidermal growth factors) have receptors expressed by endothelial cells and have been shown to have neuroprotective effects in different neurodegenerative disease models, including Parkinson’s disease.^92^ Angioneurins affect both neural and vascular cell function and regulate angiogenesis, BBB integrity, neuroregeneration, neuroprotection and synaptic plasticity.^92^

Even though statistically significant, the correlations observed with both oligodendrocytes and endothelial cells were modest, indicating a need for further studies with animal models and larger samples of Parkinson’s disease patients and later disease stages.

### Regions prone to atrophy are enriched for synaptic genes

Regions with greater atrophy progression were enriched for genes implicated in synaptic plasticity, chemical synaptic transmission, trans-synaptic signaling and cell-cell signaling. This is consistent with post-mortem evidence that α-synuclein aggregates are mostly located in synapses.^25^ Indeed, Lewy neurites, consisting of α-synuclein aggregates in presynaptic terminals, are a hallmark of Parkinson’s disease pathology,^93^ and one of the many functions of α-synuclein is to regulate synaptic homeostasis. Cell-culture experiments show that pathogenic α-synuclein fibrils first target pre-synaptic terminals and alter synaptic protein levels.^94,95^ Thus, it is likely that misfolded α-synuclein first accumulate at synapses, causing impaired neurotransmission followed by synaptic and eventual axonal and neuronal death.^26^ Pathological findings are mirrored by human imaging evidence showing that diffusion weighted MRI measures sensitive to neuritic damage are consistently abnormal in Parkinson’s disease.^49,96–98^ Finally, synaptic and neuritic death should be associated with reduced neuropil and hence tissue loss, which should be reflected in the DBM measures used here.

It has also been demonstrated that α-synuclein can be transmitted from cell to cell in a prion-like manner.^23,99^ Two findings from our work support the α-synuclein synaptic dysfunction and synaptic spreading hypothesis: (1) the regions with greater atrophy progression were enriched for genes related to synaptic activity, cell-cell communication and signaling, and (2) atrophy was dependent on connectivity.

The link to synaptic proteins may also explain the relative distribution of tissue loss along the sensory-limbic cortical hierarchy. We found that primary sensory and motor cortical areas exhibited significantly less progressive atrophy than transmodal or paralimbic cortex, in keeping with Braak’s observations.^19^ Primary areas have lower synaptic density,^100^ but they also exist at the periphery of the brain connectome,^55^ and are the most distant from limbic and default mode areas that appear to be affected early in the course of the disease. Thus, connectivity and local features may both account for the fact that primary areas are affected in the last stage of Parkinson’s disease.

### Strengths and limitations

Strengths of the study include the use of DBM, which allows the detection of both cortical and subcortical changes and has been shown to be more sensitive than VBM to subcortical volume loss in early Parkinson’s disease.^2,101^ Second, the atrophy progression measure accounts for the expected effects of sex and normal aging by using the W-score approach. Third, using a longitudinal design with the same participants at all time points limits drop-out effects that may affect brain measures derived from a different number of participants at each time point.^9,44,102^ Given the well-known clinical heterogeneity in participants with Parkinson’s disease,^103–105^ looking at the data from the same participants in a longitudinal study ensures greater consistency and reduces confounding factors. Fourth, having multiple follow-up time points (one, two and four years) also gives a more precise estimate of the trajectory of atrophy progression through time.

Some limitations should also be acknowledged. Follow-up duration for the clinical and neuroimaging data is relatively short (4 years) and longer follow-up should help to determine the relationships between atrophy progression and cognitive dysfunction, mood symptoms and CSF biomarkers. Moreover, the PPMI database includes measures from multiple centers which may lead to site-specific biases. However, PPMI has strict guidelines and protocols to acquire the clinical and imaging data to ensure standardization^42^ and scanner site was used as covariate in all our analyses. Finally, only participants with Parkinson’s disease with neuroimaging data at all four points were included in the main analysis, possibly excluding those with more severe symptoms and/or more atrophy (who are more predisposed to drop out of the study) and raising the possibility of survivor and collider bias.^45^ The former could mask disease progression, while the latter could lead to biased estimates of associations. Note however that additional analyses were performed with larger sample sizes including participants with data at baseline and one year, and baseline and two years, and results were comparable. Finally, other structural neuroimaging measures (such as cortical thickness or diffusion-weighted MRI) should also be used to investigate the longitudinal progression of atrophy.

## Conclusion

Widespread atrophy was found after four years in the early stages of Parkinson’s disease. This pattern identified vulnerable brain regions that could eventually be used as a guide for other Parkinson’s disease longitudinal studies. We also showed factors associated with disease progression, including connectivity and regional tissue composition. This may help identify biological processes implicated in the progression of Parkinson’s disease. In addition, our results provide further support for the α-synuclein network spread hypothesis first proposed by Braak.

## Acknowledgements

We would like to thank Jakob Seidlitz for sharing his cell-type gene sets and for his valuable advice. We also thank Yashar Zeighami and Alexandre Hutton for their assistance. Data used in this article were obtained from the PPMI database (www.ppmi-info.org/data). For up-to-date information on the study, visit: www.ppmi-info.org. PPMI, a public-private partnership, funded by the Michael J. Fox Foundation for Parkinson’s Research and funding partners, including AbbVie, Avid Radiopharmaceuticals, Biogen, Bristol-Myers Squibb, Covance, GE Healthcare, Genentech, GlaxoSmithKline (GSK), Eli Lilly and Company, Lundbeck, Merck, Meso Scale Discovery (MSD), Pfizer, Piramal Imaging, Roche, Sanofi Genzyme, Servier, Teva, and UCB (www.ppmi-info.org/fundingpartners).

## Supplementary material

Supplementary material is available online.

## Abbreviations

AHBA: Allen Human Brain Atlas
BBB: blood-brain barrier
Bl: Baseline
CAT12: Computational Anatomy Toolbox
DBM: deformation-based morphometry
FC: functional connectivity
FDR: false discovery rate
FEW: Family-wise error
GDS: Geriatric Depression Scale
GO: gene ontology
GOrilla: Gene Ontology enRIchment anaLysis and visuaLizAtion tool
HC: healthy control
HVLT: Hopkins Verbal Learning Test
L-Dopa: Levodopa
MDS-UPDRS: Movement Disorder Society Unified Parkinson Disease Rating Scale
MNI: Montreal Neurological Institute
MoCA: Montreal Cognitive Assessment
NFL: neurofilament light polypeptide
PPMI: Parkinson’s progression markers initiative
pTFCE: probabilistic approach for threshold-free cluster enhancement
SD: Standard Deviation
SPM12: Statistical Parametric Mapping software
STAI: State-Trait Anxiety Inventory

