## Supplementary Material for "Brain atrophy progression in Parkinson’s disease is shaped by connectivity and local vulnerability"

### *Supplemental Information*

**Inclusion criteria (participants with Parkinson's disease)**

(1) at least two of the following: resting tremor, bradykinesia, and rigidity *or* either asymmetric resting tremor *or* asymmetric bradykinesia

(2) a diagnosis of Parkinson's disease for 2 years or less at the screening stage

(3) Hoehn & Yahr stage I or II at the baseline visit

(4) confirmation of dopamine transporter deficit by single-photon emission computed tomography

(5) not expected to require Parkinson's disease medications within at least 6 months of the baseline visit

(6) at least 30 years old at the time of Parkinson's disease diagnosis

**Deformation based morphometry (DBM)**

In brief, the raw T1-weighted MRI images were processed through an initial processing during which the image was resampled, corrected for bias field inhomogeneities, affine-registered, and entered into SPM's unified segmentation protocol. Next, the image underwent a more refined processing that includes skull stripping, brain parcellation, local tissue intensity transformation, partial volume estimation, and spatial normalization to a common reference using DARTEL. For each participant at each time point, the DBM pipeline produced a set of processed images including a voxel-wise map of the Jacobian determinants, which was used as a marker of local brain atrophy. A 2-mm FWHM smoothing kernel was then applied on the Jacobian maps and the log of the Jacobian determinant was calculated as the voxel-wise relative deformation value to create DBM maps. DBM processing was carried out with the Computational Anatomy Toolbox (CAT12)[1] implemented in Statistical Parametric Mapping software (SPM12, https://www.fil.ion.ucl.ac.uk/spm/doc/).

**Structural and Functional Network Analysis**

*1) Regional brain atrophy progression*

The brain was parcellated into 68, 114, 219, and 448 approximately equally sized cortical regions using the Cammoun atlas.[2] Each analysis was repeated at all four resolutions to ensure that the results were robust regardless of the spatial scale. The average atrophy progression (i.e., mean W-Score difference between the follow-up time point and baseline) was calculated for each cortical region.

*2) Structural and functional network construction*

Structural and functional connectivity matrices were respectively constructed from diffusion-weighted and resting-state functional MRI scans acquired from 70 healthy adults (age = 29 ± 9 year, 43 men)[3] (https://doi.org/10.5281/zenodo.2872624). For the structural connectome, a deterministic streamline tractography approach was used. A binary group-average structural connectivity matrix was computed for each of the four resolution scales using a distance-dependent consensus approach that preserved the connection-length distribution of each participant[4] (https://www.brainnetworkslab.com/coderesources). In addition, the mean functional connectivity of all the 70 participants was calculated to obtain the group-average functional connectome.

*3) Collective neighborhood atrophy progression*

The group-level structural and functional connectivity matrices were used to compute the collective atrophy progression of the neighborhood of each region, an index of each node's "disease exposure". The collective atrophy progression of structurally connected neighbors ($D_i$) was calculated as the mean atrophy progression of all the regions with a (deterministic) structural connection with a given region $i$ (node) (see Equation 1). The collective atrophy progression of functional neighbors was calculated in a similar way, but with each neighbor $j$'s deformation value being weighted by the strength of the functional connectivity (FC) between node $i$ and $j$ (see Equation 2).

$$D_i = \frac{1}{N_i} \sum_{i \neq j, j=1}^{N_i} d_j \quad (1) \qquad\qquad D_i = \frac{1}{N_i} \sum_{i \neq j, j=1}^{N_i} d_j \times FC_{ij} \quad (2)$$

In both equations, $D_i$ is the disease exposure or average deformation progression in the neighbors of region $i$, $d$ is the region atrophy progression, and $N_i$ is the number of regions structurally connected to the region $i$ (i.e., node degree). Therefore, only nodes with a direct structural connection are assumed to contribute to each other's atrophy progression. The deformation in the region under consideration was excluded from equations (1) and (2) i.e. regions did not contribute to their own atrophy progression. For the functional neighborhood atrophy progression (equation 2), only the regions structurally connected to the given region were considered.

*4) Correlations*

Pearson's correlations were calculated between the mean atrophy progression of all brain regions and the collective atrophy progression of their structurally connected neighbors and their structurally connected neighbors weighted by their FC.

*5) Comparison with a null model*

To investigate whether the relationships between the atrophy progression of a region and the atrophy progression of its structurally and functionally connected neighborhood depends on the network spatial topology, the correlations were tested against a null model preserving the spatial autocorrelation between regions[5] using the netneurotools toolbox (https://netneurotools.readthedocs.io/en/latest/index.html). This approach generates a null model by projecting the brain regions onto a sphere and randomly rotating the sphere. To address the loss of data caused by the medial wall rotation, this model assigns the nearest data to a missing parcel (see Markello and

Misic[6]) for a comparison between spatially-constrained null models). Briefly, a surface-based representation of the different resolution of the Cammoun atlas on the FreeSurfer (release v6.0.0; http://surfer.nmr.mgh.harvard.edu/) fsaverage surface was created. The spherical projection of the fsaverage surface was then used to define spatial coordinates for each region by selecting the vertex closest to the center of mass of the region. The spatial coordinates were used to create null models by rotating and reassigning region values 10,000 times. The regions centroids were assigned to the closest rotated centroids based on the minimum Euclidean distance. The spatial rotation was performed at the parcel resolution and in one hemisphere before being mirrored in the other. Finally, Pearson's correlation coefficients that were derived from the empirical networks (i.e., observed values) were tested against those derived from the null networks (i.e., permuted values).

*References*

**Table 1.** Regions showing a significant atrophy progression with time (F-Contrast)

| F-Value | Cluster Size Number of voxels | Peak xyz [mm] | Overlap with the neuromorphometric atlas | Brain regions |
|---|---|---|---|---|
| 25.34 | 24366 | 57 -4 -16 | 41.00% | Right Cerebral White Matter |
| | | | 14.00% | Left Cerebral White Matter |
| | | | 6.00% | Right MTG middle temporal gyrus |
| | | | 5.00% | Right AnG angular gyrus |
| | | | 4.00% | Right PCu precuneus |
| | | | 4.00% | Left Lateral Ventricle |
| | | | 3.00% | Right TMP temporal pole |
| | | | 3.00% | Right Lateral Ventricle |
| | | | 3.00% | Right ITG inferior temporal gyrus |
| | | | 3.00% | Unknown |
| | | | 2.00% | Right FuG fusiform gyrus |
| | | | 2.00% | Right SPL superior parietal lobule |
| | | | 2.00% | Right STG superior temporal gyrus |
| | | | 2.00% | Right PCgG posterior cingulate gyrus |
| | | | 1.00% | Right MOG middle occipital gyrus |
| | | | 1.00% | Right IOG inferior occipital gyrus |
| 21.33 | 11517 | -52 -46 8 | 35.00% | Left Cerebral White Matter |
| | | | 19.00% | Left MTG middle temporal gyrus |
| | | | 8.00% | Left TMP temporal pole |
| | | | 6.00% | Left ITG inferior temporal gyrus |
| | | | 6.00% | Left STG superior temporal gyrus |
| | | | 5.00% | Left AnG angular gyrus |
| | | | 5.00% | Left PCu precuneus |
| | | | 3.00% | Left SPL superior parietal lobule |
| | | | 3.00% | Left MOG middle occipital gyrus |
| | | | 2.00% | Left FuG fusiform gyrus |
| | | | 2.00% | Left IOG inferior occipital gyrus |
| | | | 1.00% | Left PCgG posterior cingulate gyrus |
| | | | 1.00% | Left SMG supramarginal gyrus |
| | | | 1.00% | Left Amygdala |
| | | | 1.00% | Left Ent entorhinal area |
| 20.6 | 2396 | 12 -32 -40 | 66.00% | Brain Stem |
| | | | 17.00% | Left Cerebellum White Matter |
| | | | 17.00% | Right Cerebellum White Matter |
| 16.18 | 388 | -46 -26 44 | 37.00% | Left Cerebral White Matter |
| | | | 34.00% | Left PoG postcentral gyrus |
| | | | 25.00% | Left SMG supramarginal gyrus |
| | | | 4.00% | Left PO parietal operculum |
| 21.16 | 286 | -14 12 58 | 54.00% | Left Cerebral White Matter |
| | | | 46.00% | Left SFG superior frontal gyrus |
| 17.4 | 159 | -8 20 -12 | 44.00% | Left Cerebral White Matter |
| | | | 20.00% | Left SCA subcallosal area |
| | | | 19.00% | Left MFC medial frontal cortex |
| | | | 14.00% | Left ACgG anterior cingulate gyrus |
| | | | 3.00% | Left Accumbens Area |
| 15.74 | 116 | 14 16 0 | 97.00% | Right Caudate |
| | | | 3.00% | Right Cerebral White Matter |
| 19.74 | 61 | 14 -74 -6 | 52.00% | Right LiG lingual gyrus |
| | | | 48.00% | Right Cerebral White Matter |
| 16.78 | 88 | -40 21 12 | 88.00% | Left Cerebral White Matter |
| | | | 9.00% | Left TrIFG triangular part of the inferior frontal gyrus |
| | | | 3.00% | Left OpIFG opercular part of the inferior frontal gyrus |
| 15.34 | 59 | 16 26 16 | 100.00% | Right Cerebral White Matter |
| 15.26 | 60 | 8 28 32 | 98.00% | Right MSFG superior frontal gyrus medial segment |
| | | | 2.00% | Right ACgG anterior cingulate gyrus |
| 15.16 | 59 | 4 16 -18 | 56.00% | Right Cerebral White Matter |
| | | | 31.00% | Right SCA subcallosal area |
| | | | 14.00% | Right Accumbens Area |
| 14.83 | 13 | -46 -20 20 | 100.00% | Left CO central operculum |
| 14.45 | 79 | 22 -9 -21 | 72.00% | Right Hippocampus |
| | | | 28.00% | Right Amygdala |
| 13.89 | 99 | 42 -34 44 | 74.00% | Right SMG supramarginal gyrus |
| | | | 14.00% | Right Cerebral White Matter |
| | | | 7.00% | Right PoG postcentral gyrus |
| | | | 5.00% | Right SPL superior parietal lobule |
| 13.7 | 16 | 16 33 3 | 100.00% | Right Cerebral White Matter |
| 13.69 | 18 | 16 12 46 | 100.00% | Right Cerebral White Matter |
| 13.67 | 10 | -3 20 64 | 70.00% | Left SMC supplementary motor cortex |
| | | | 20.00% | Left SFG superior frontal gyrus |
| | | | 10.00% | Unknown |
| 13.66 | 14 | 30 -9 -32 | 79.00% | Right PHG parahippocampal gyrus |
| | | | 14.00% | Right Cerebral White Matter |
| | | | 7.00% | Right FuG fusiform gyrus |
| 13.55 | 11 | 2 14 64 | 91.00% | Unknown |
| | | | 9.00% | Left SMC supplementary motor cortex |
| 13.46 | 13 | -4 9 38 | 100.00% | Left MCgG middle cingulate gyrus |
| 13.29 | 15 | -2 39 44 | 100.00% | Left MSFG superior frontal gyrus medial segment |
| 13.28 | 12 | 12 12 58 | 100.00% | Right Cerebral White Matter |
| 13.09 | 11 | 26 4 -30 | 55.00% | Right TMP temporal pole |
| | | | 45.00% | Right Ent entorhinal area |

**Table 2.** Regions showing a significant atrophy progression after two and four years (T-Contrasts)

| T-Value | Cluster Size Number of voxels | Peak xyz [mm] | Overlap with the neuromorphometric atlas | Brain regions |
|---|---|---|---|---|
| | | | **Contrast: Baseline - 2 years** | |
| 6.55 | 487 | 46 -62 12 | 40.00% | Right AnG angular gyrus |
| | | 45 -62 26 | 31.00% | Right Cerebral White Matter |
| | | | 25.00% | Right MTG middle temporal gyrus |
| 6.68 | 285 | 57 -4 -16 | 53.00% | Right MTG middle temporal gyrus |
| | | 57 -15 -10 | 43.00% | Right STG superior temporal gyrus |
| | | | 5.00% | Right Cerebral White Matter |
| 6.66 | 188 | 14 15 2 | 94.00% | Right Caudate |
| 5.98 | 143 | -57 -28 -22 | 62.00% | Left MTG middle temporal gyrus |
| | | | 20.00% | Left Cerebral White Matter |
| | | | 19.00% | Left ITG inferior temporal gyrus |
| 6.16 | 69 | -52 3 -22 | 51.00% | Left TMP temporal pole |
| | | | 36.00% | Left MTG middle temporal gyrus |
| 6.94 | 43 | -6 -54 38 | 98.00% | Left PCu precuneus |
| 5.77 | 27 | 33 9 -42 | 100.00% | Right TMP temporal pole |
| 5.69 | 18 | 46 12 -33 | 100.00% | Right TMP temporal pole |
| 5.78 | 14 | -56 -6 -10 | 100.00% | Left STG superior temporal gyrus |
| 5.73 | 13 | -56 -45 9 | 100.00% | Left STG superior temporal gyrus |
| 5.65 | 10 | -60 -15 -12 | 100.00% | Left MTG middle temporal gyrus |
| | | | **Contrast: Baseline - 4 years** | |
| 8.53 | 48093 | 40 -64 39 | 22.00% | Right Cerebral White Matter |
| | | | 18.00% | Left Cerebral White Matter |
| | | | 7.00% | Left MTG middle temporal gyrus |
| | | | 5.00% | Right MTG middle temporal gyrus |
| | | | 4.00% | Right PCu precuneus |
| | | | 3.00% | Right AnG angular gyrus |
| | | | 3.00% | Left ITG inferior temporal gyrus |
| | | | 3.00% | Left TMP temporal pole |
| | | | 3.00% | Right TMP temporal pole |
| | | | 2.00% | Left PCu precuneus |
| | | | 2.00% | Left AnG angular gyrus |
| | | | 2.00% | Left STG superior temporal gyrus |
| | | | 2.00% | Right ITG inferior temporal gyrus |
| | | | 2.00% | Right SPL superior parietal lobule |
| | | | 2.00% | Right FuG fusiform gyrus |
| | | | 1.00% | Left IOG inferior occipital gyrus |
| | | | 1.00% | Right STG superior temporal gyrus |
| | | | 1.00% | Left MOG middle occipital gyrus |
| | | | 1.00% | Left SPL superior parietal lobule |
| | | | 1.00% | Right MOG middle occipital gyrus |
| | | | 1.00% | Right PCgG posterior cingulate gyrus |
| | | | 1.00% | Right SMG supramarginal gyrus |
| 7.63 | 234 | 14 -74 -6 | 58.00% | Right LiG lingual gyrus |
| | | | 42.00% | Right Cerebral White Matter |
| 7.42 | 286 | -14 12 58 | 50.00% | Left Cerebral White Matter |
| | | | 50.00% | Left SFG superior frontal gyrus |
| 6.81 | 131 | -40 21 12 | 94.00% | Left Cerebral White Matter |
| | | | 5.00% | Left OpIFG opercular part of the inferior frontal gyrus |
| 6.73 | 239 | -9 21 -12 | 39.00% | Left Cerebral White Matter |
| | | | 21.00% | Left SCA subcallosal area |
| | | | 19.00% | Left MFC medial frontal cortex |
| | | | 18.00% | Left ACgG anterior cingulate gyrus |
| | | | 2.00% | Left Accumbens Area |
| 6.14 | 47 | 6 38 27 | 100.00% | Right MSFG superior frontal gyrus medial segment |
| 6.13 | 296 | 15 20 -4 | 58.00% | Right Caudate |
| | | | 23.00% | Right Cerebral White Matter |
| | | | 10.00% | Right Accumbens Area |
| | | | 9.00% | Right SCA subcallosal area |
| 5.76 | 106 | -33 -45 42 | 63.00% | Left SPL superior parietal lobule |
| | | | 25.00% | Left SMG supramarginal gyrus |
| | | | 11.00% | Left Cerebral White Matter |
| 5.53 | 16 | 58 -3 -36 | 100.00% | Unknown |
| 5.48 | 55 | 20 -84 -12 | 49.00% | Right LiG lingual gyrus |
| | | | 35.00% | Right Cerebral White Matter |
| | | | 16.00% | Right OFuG occipital fusiform gyrus |
| 5.46 | 15 | -14 -44 70 | 60.00% | Left PoG postcentral gyrus |
| | | | 40.00% | Left Cerebral White Matter |
| 5.43 | 14 | 15 -78 -14 | 100.00% | Right LiG lingual gyrus |
| 5.42 | 10 | 20 -98 -9 | 60.00% | Right OCP occipital pole |
| | | | 20.00% | Right Cerebral White Matter |
| | | | 20.00% | Right OFuG occipital fusiform gyrus |

**Table 3.** Regions showing a significant atrophy progression after one and two years after including all the subjects with a T1-MRI at baseline and one year (*N*=120), and baseline and two years (*N*=109)

| T-Value | Cluster Size Number of voxels | Peak xyz [mm] | Overlap with the neuromorphometric atlas | Brain regions |
|---|---|---|---|---|
| | | **Contrast: Baseline - 1 year** | | |
| 6.38 | 58 | -63 -32 -4 | 98.00% | Left MTG middle temporal gyrus |
| | | | 2.00% | Left Cerebral White Matter |
| 6.37 | 42 | -52 -18 -30 | 86.00% | Left ITG inferior temporal gyrus |
| | | | 14.00% | Left Cerebral White Matter |
| 6.24 | 89 | -58 -16 -14 | 92.00% | Left MTG middle temporal gyrus |
| | | | 6.00% | Left Cerebral White Matter |
| | | | 2.00% | Left STG superior temporal gyrus |
| 6.13 | 20 | 18 18 4 | 95.00% | Right Caudate |
| | | | 5.00% | Right Cerebral White Matter |
| 6.01 | 40 | 10 8 10 | 100.00% | Right Caudate |
| 5.97 | 53 | -56 -48 -4 | 51.00% | Left MTG middle temporal gyrus |
| | | | 49.00% | Left Cerebral White Matter |
| 5.94 | 20 | -45 -30 -21 | 90.00% | Left ITG inferior temporal gyrus |
| | | | 5.00% | Left Cerebral White Matter |
| | | | 5.00% | Left FuG fusiform gyrus |
| 5.77 | 17 | -56 -28 -22 | 59.00% | Left ITG inferior temporal gyrus |
| | | | 41.00% | Left Cerebral White Matter |
| 5.74 | 13 | -54 -3 -26 | 62.00% | Left MTG middle temporal gyrus |
| | | | 38.00% | Left Cerebral White Matter |
| | | **Contrast: Baseline -2 years** | | |
| 8.71 | 4530 | -56 -27 -22 | 34.00% | Left MTG middle temporal gyrus |
| | | | 19.00% | Left TMP temporal pole |
| | | | 15.00% | Left Cerebral White Matter |
| | | | 11.00% | Left STG superior temporal gyrus |
| | | | 10.00% | Left ITG inferior temporal gyrus |
| | | | 5.00% | Left AnG angular gyrus |
| | | | 4.00% | Left FuG fusiform gyrus |
| 7.62 | 817 | 57 -3 -18 | 37.00% | Right TMP temporal pole |
| | | | 36.00% | Right MTG middle temporal gyrus |
| | | | 16.00% | Right STG superior temporal gyrus |
| | | | 8.00% | Right Cerebral White Matter |
| | | | 2.00% | Right ITG inferior temporal gyrus |
| 7.61 | 655 | 15 21 -3 | 87.00% | Right Caudate |
| | | | 6.00% | Right Cerebral White Matter |
| | | | 6.00% | Right Accumbens Area |
| 7.55 | 823 | 46 -60 12 | 46.00% | Right Cerebral White Matter |
| | | | 27.00% | Right MTG middle temporal gyrus |
| | | | 23.00% | Right AnG angular gyrus |
| | | | 3.00% | Right MOG middle occipital gyrus |
| 6.68 | 129 | -50 24 6 | 74.00% | Left Cerebral White Matter |
| | | | 13.00% | Left OpIFG opercular part of the inferior frontal gyrus |
| | | | 13.00% | Left TrIFG triangular part of the inferior frontal gyrus |
| 6.28 | 22 | 58 -3 -36 | 86.00% | Unknown |
| | | | 14.00% | Right ITG inferior temporal gyrus |
| 6.27 | 39 | 26 -68 36 | 72.00% | Right SPL superior parietal lobule |
| | | | 13.00% | Right Cerebral White Matter |
| | | | 8.00% | Right AnG angular gyrus |
| | | | 8.00% | Right SOG superior occipital gyrus |
| 6.19 | 14 | 4 -51 24 | 64.00% | Right PCgG posterior cingulate gyrus |
| | | | 36.00% | Right PCu precuneus |
| 6.04 | 26 | -6 -56 38 | 100.00% | Left PCu precuneus |
| 5.76 | 12 | 28 -8 -33 | 50.00% | Right PHG parahippocampal gyrus |
| | | | 42.00% | Right FuG fusiform gyrus |
| | | | 8.00% | Right Cerebral White Matter |
| 5.75 | 14 | 44 15 -26 | 57.00% | Right TMP temporal pole |
| | | | 43.00% | Right Cerebral White Matter |
| 5.74 | 36 | 39 -57 28 | 67.00% | Right Cerebral White Matter |
| | | | 33.00% | Right AnG angular gyrus |

**Figure 1.** Overlap between the regions showing significant atrophy progression ($P_{FWE}$-*value*<0.05) after including the subjects with a T1-MRI at all time points (*N*=74, Bl-2Y: red) and all the subjects with a T1-MRI at baseline and one year (*N*=120, Bl-1Y: green), and baseline and two years (*N*=109, Bl-2Y: blue) using the MNI152-2009c template for visualization purpose (Top row: $x$ = 45, $y$ = -62, $z$ = -19; Middle row: $x$ = 14, $y$ = 15, $z$ = 7; Bottom row: $x$ = 54, $y$ = -6, $z$ = 30). P: posterior; A: anterior; R: right; L: left

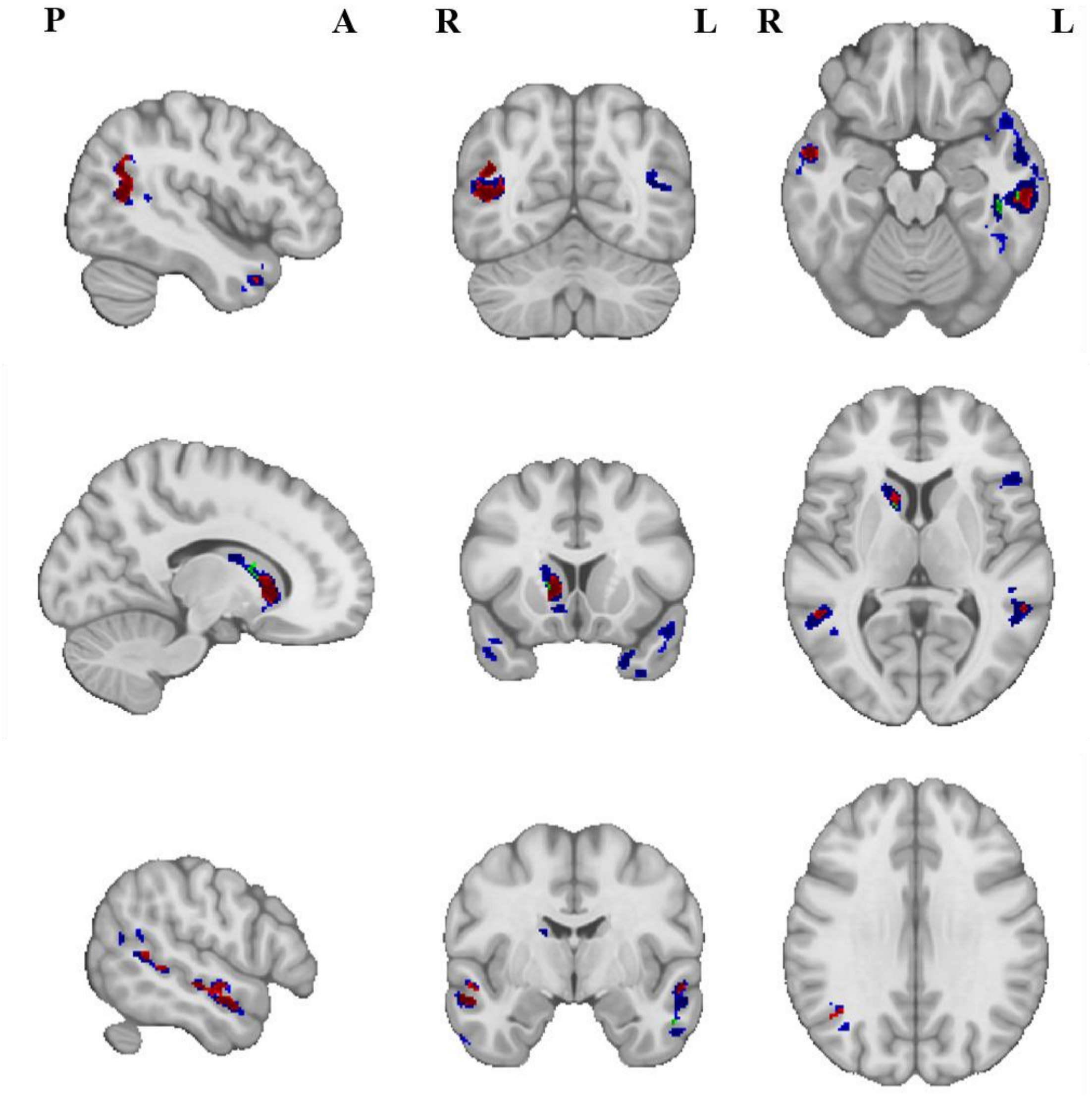

**Figure 2.** Relationship between the regional atrophy progression and the atrophy progression in their structurally (SC) and functionally connected (FC) neighbors after two and four years using the Cammoun atlas with 68 (I), 114 (II) and 219 (III) regions.

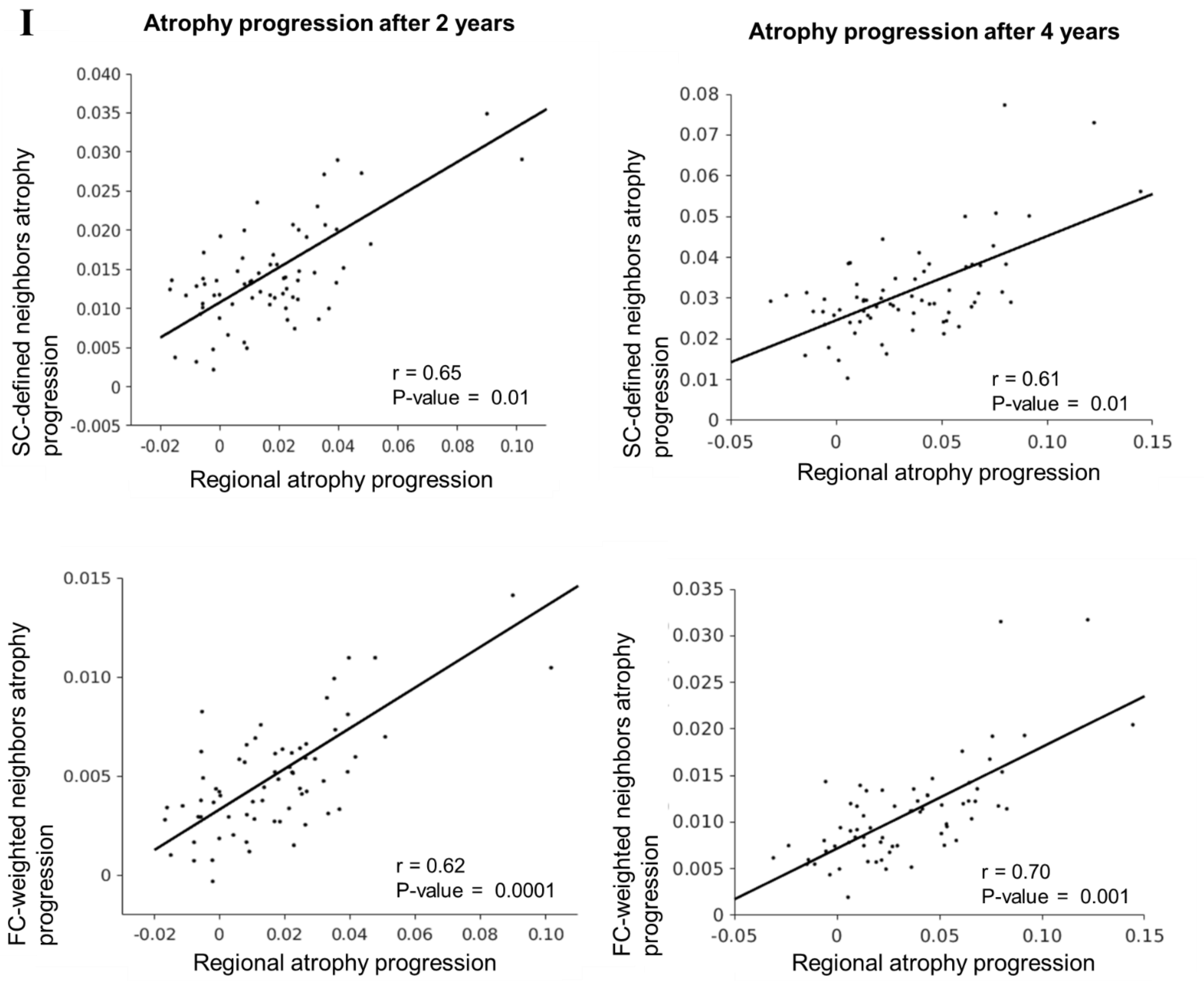

**II**

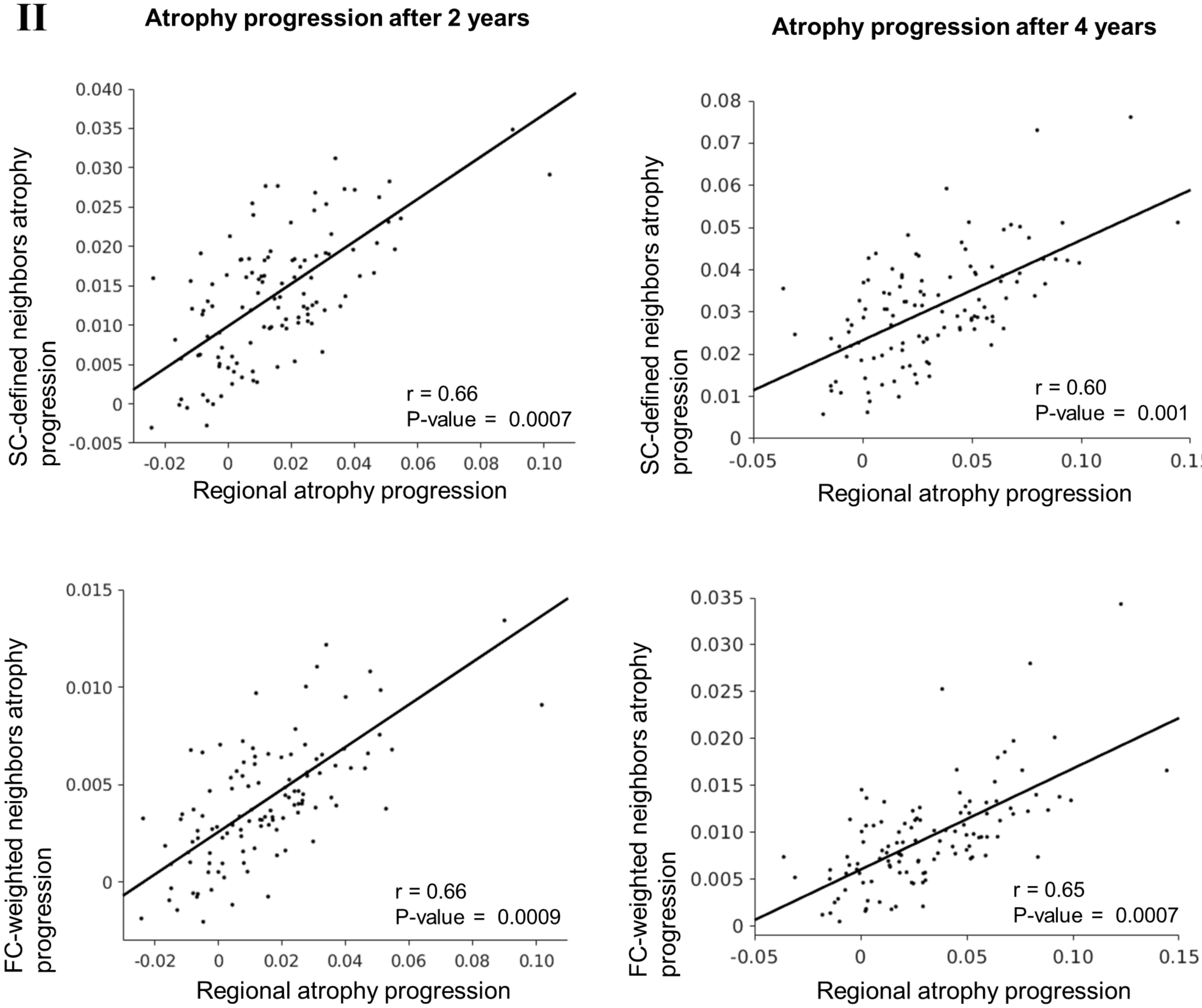
Atrophy progression after 2 years
Atrophy progression after 4 years
SC-defined neighbors atrophy progression
FC-weighted neighbors atrophy progression
Regional atrophy progression
r = 0.66
P-value = 0.0007
r = 0.60
P-value = 0.001
r = 0.66
P-value = 0.0009
r = 0.65
P-value = 0.0007

**III**

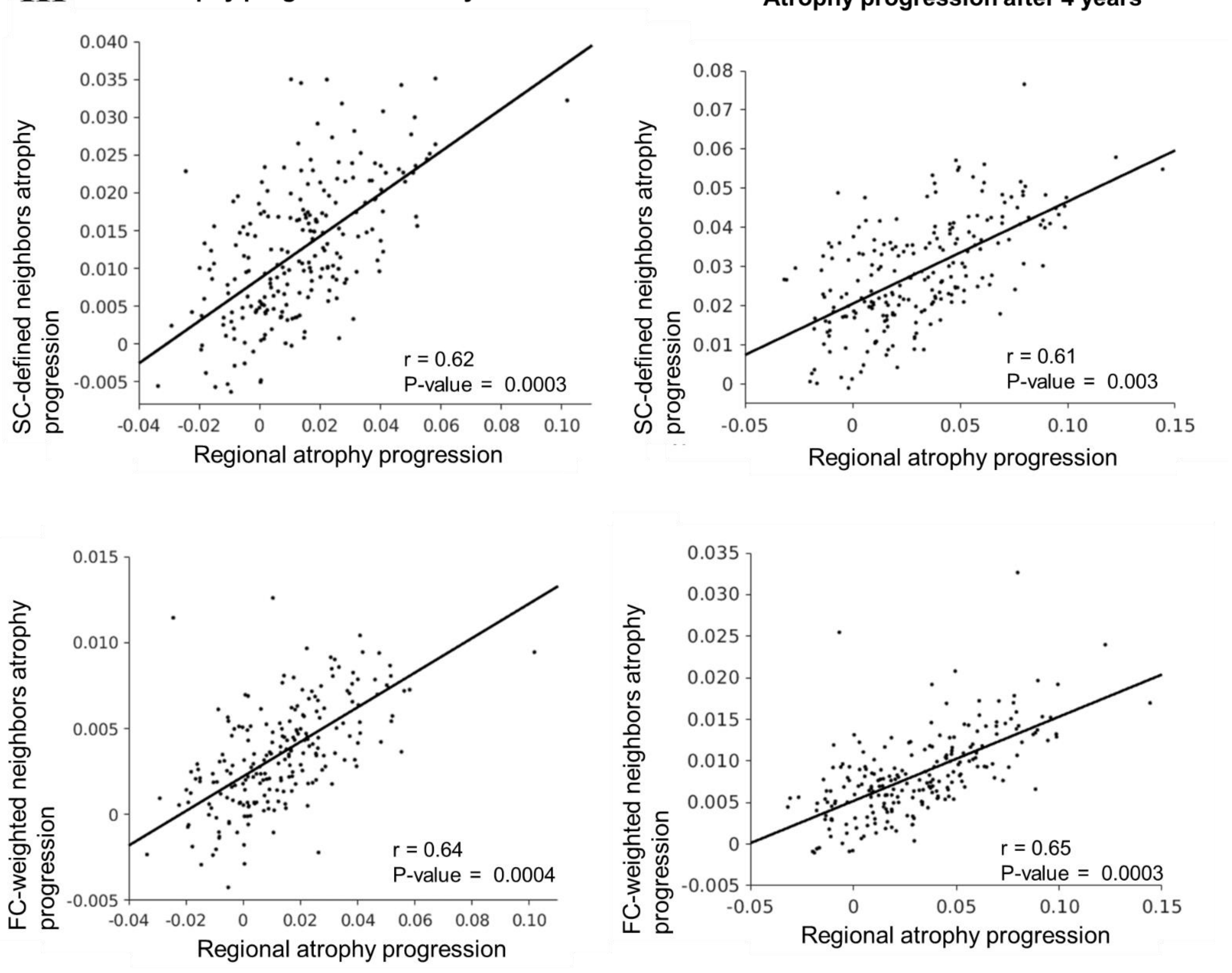

Atrophy progression after 2 years
Atrophy progression after 4 years
SC-defined neighbors atrophy progression
FC-weighted neighbors atrophy progression
Regional atrophy progression
r = 0.62
P-value = 0.0003
r = 0.61
P-value = 0.003
r = 0.64
P-value = 0.0004
r = 0.65
P-value = 0.0003

**Figure 3.** Relationship between the regional atrophy progression after two and four years and the prevalence of endothelial cells and oligodendrocytes using the Cammoun atlas with 68 (I), 114 (II) and 219 (III) regions.

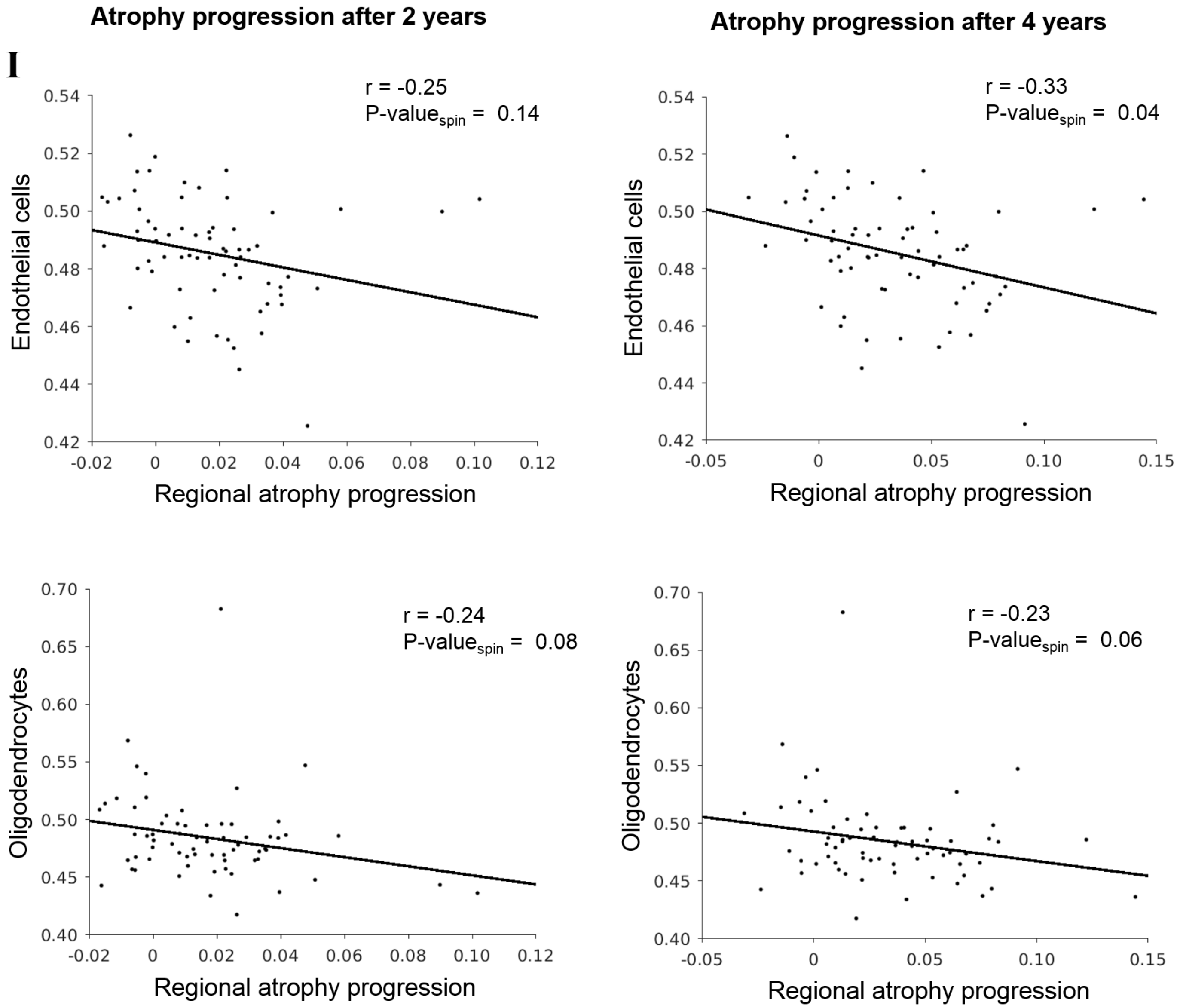

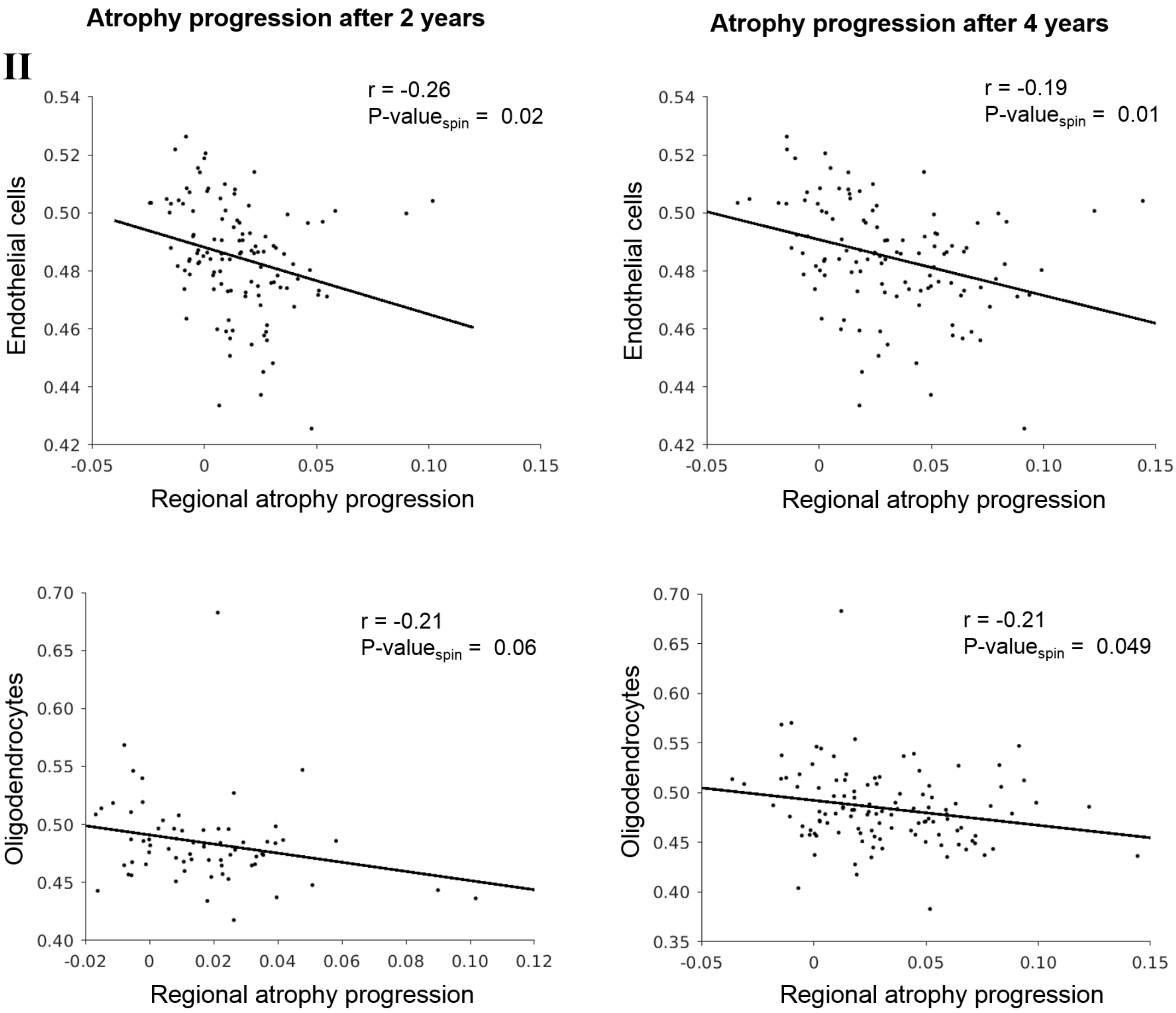
Atrophy progression after 2 years
Atrophy progression after 4 years
II
r = -0.26
P-value$_{spin}$ = 0.02
r = -0.19
P-value$_{spin}$ = 0.01
Endothelial cells
Regional atrophy progression
r = -0.21
P-value$_{spin}$ = 0.06
r = -0.21
P-value$_{spin}$ = 0.049
Oligodendrocytes
Regional atrophy progression

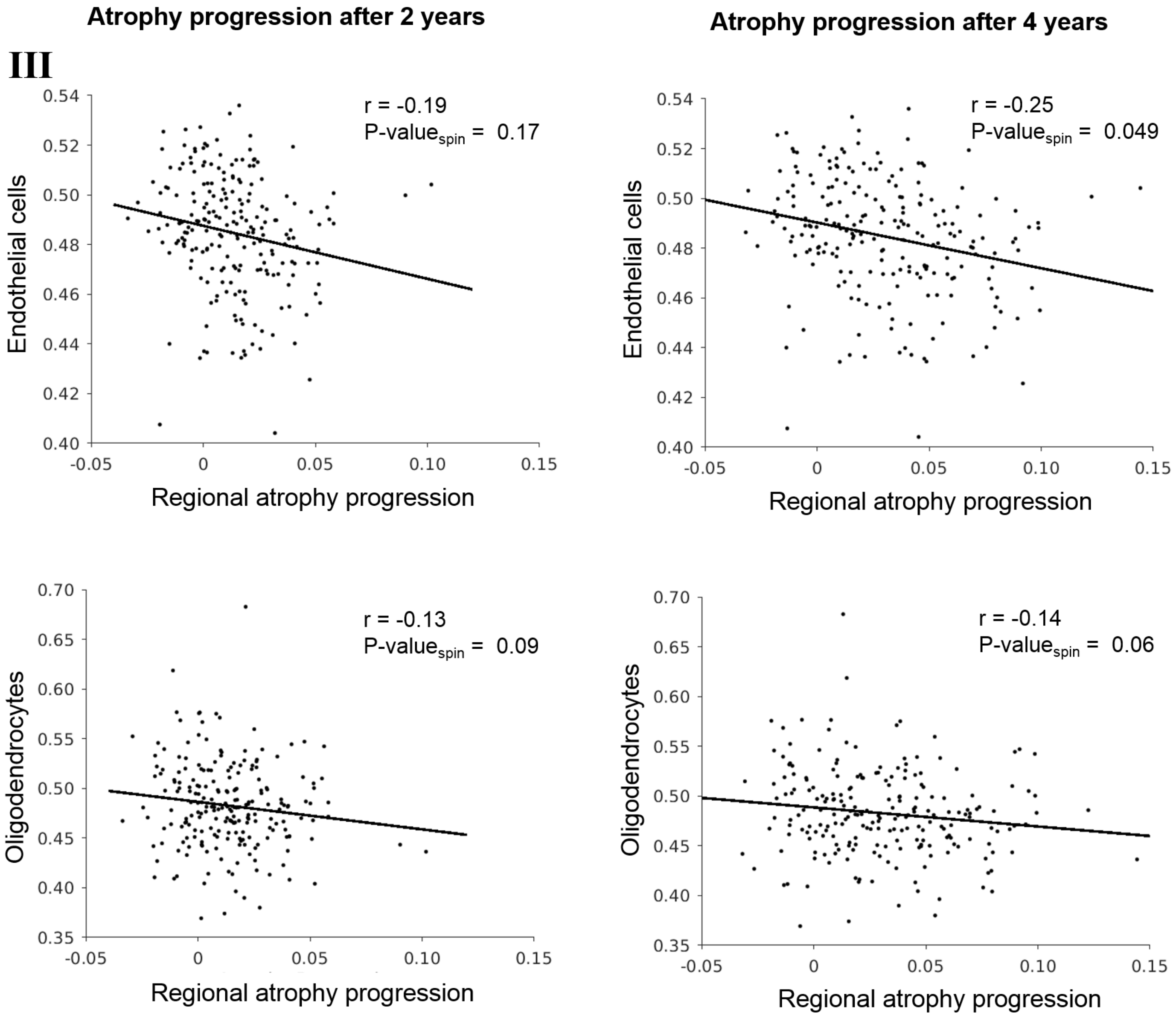
Atrophy progression after 2 years
Atrophy progression after 4 years
III
r = -0.19
P-value$_{spin}$ = 0.17
r = -0.25
P-value$_{spin}$ = 0.049
Endothelial cells
Regional atrophy progression
r = -0.13
P-value$_{spin}$ = 0.09
r = -0.14
P-value$_{spin}$ = 0.06
Oligodendrocytes
Regional atrophy progression

**Figure 4.** No significant correlation between the atrophy progression after two and four years in *de novo* Parkinson’s disease and the prevalence of astrocytes (I), microglia (II), oligodendrocyte precursors cells (III), excitatory neurons (IV) and inhibitory neurons (V) in the cortex.

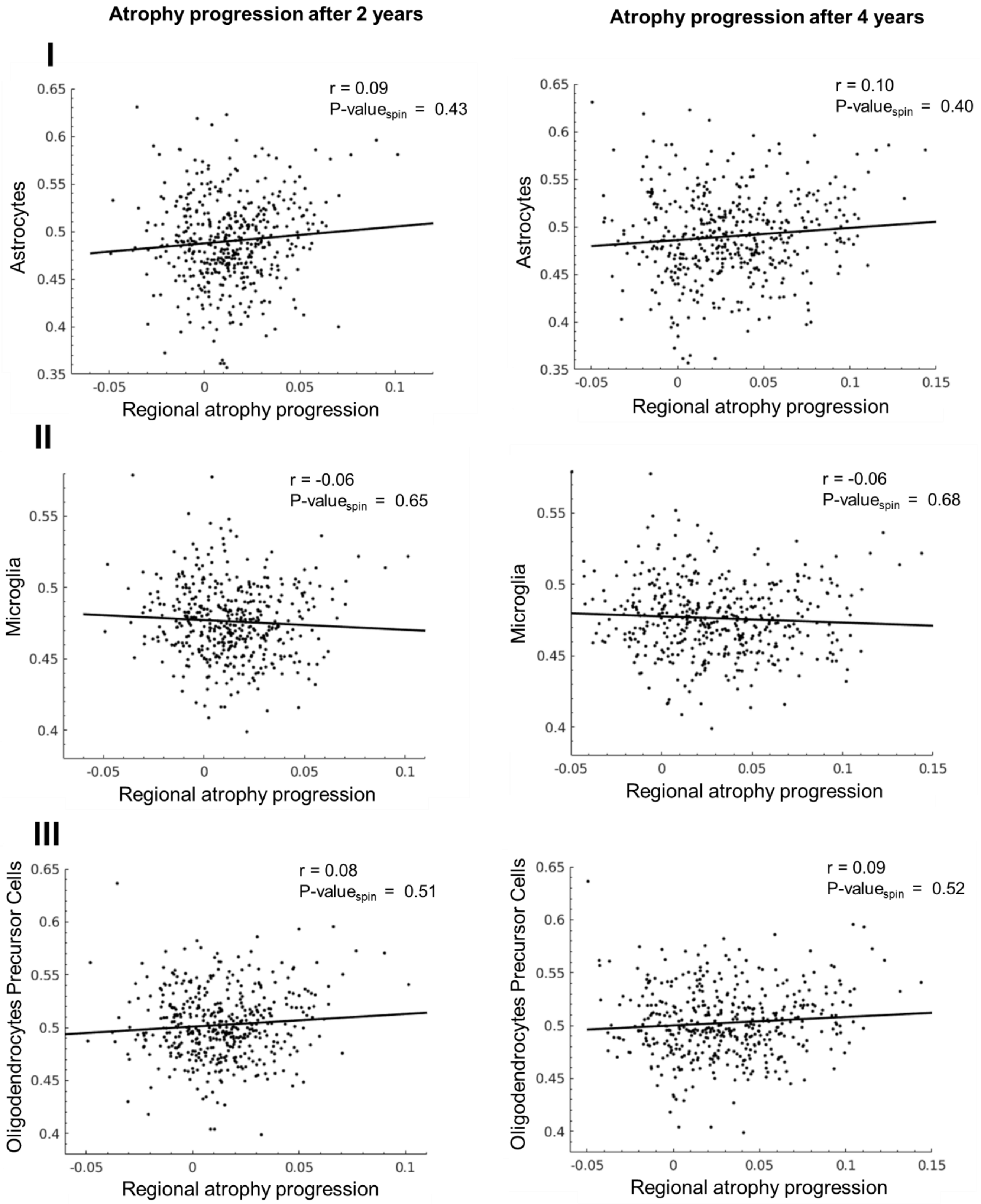

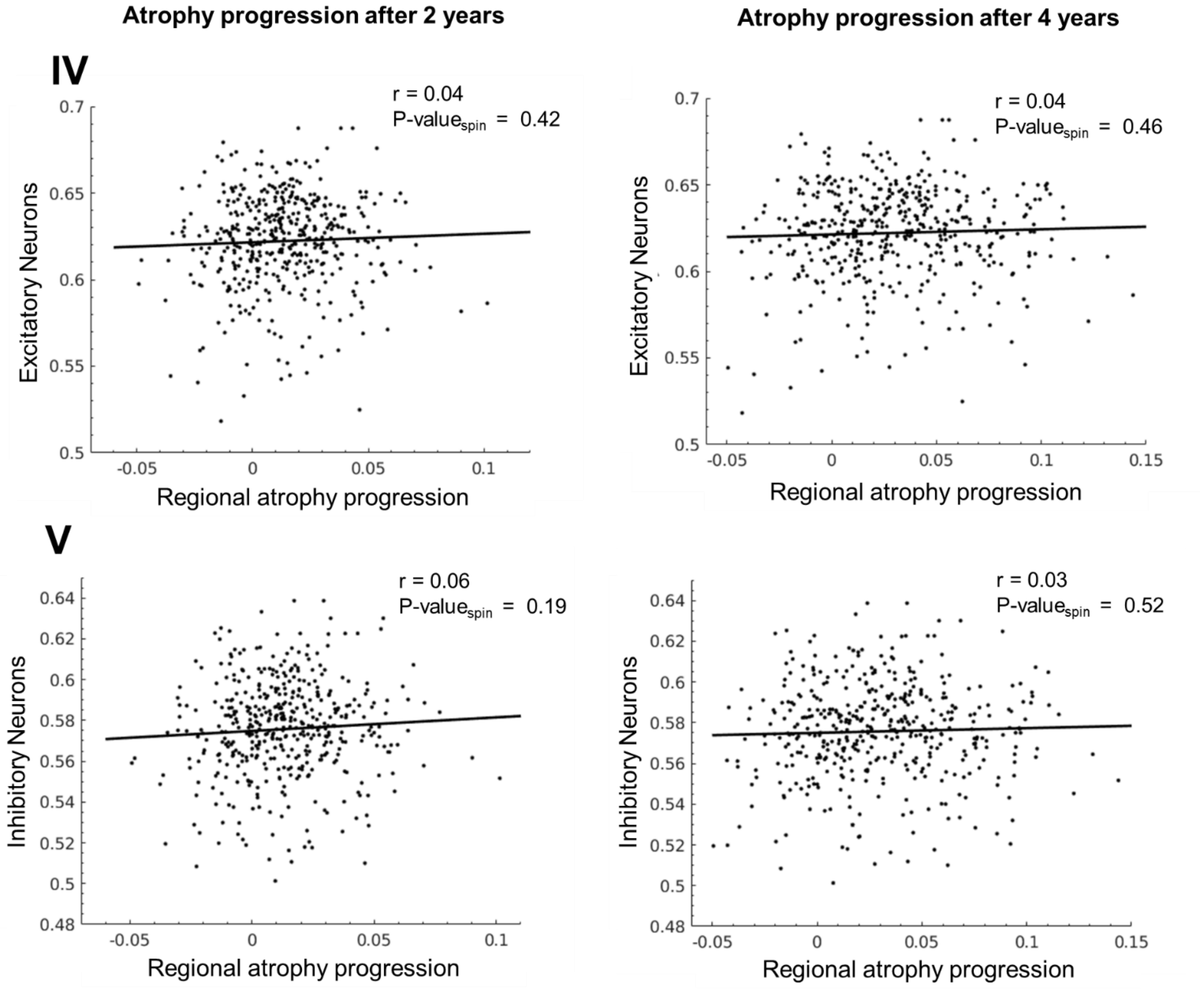
Atrophy progression after 2 years
Atrophy progression after 4 years
IV
V
r = 0.04
P-value$_{spin}$ = 0.42
r = 0.04
P-value$_{spin}$ = 0.46
r = 0.06
P-value$_{spin}$ = 0.19
r = 0.03
P-value$_{spin}$ = 0.52
Excitatory Neurons
Inhibitory Neurons
Regional atrophy progression

**Figure 5.** Gene ontology (GO) enrichment analysis of all the genes related to atrophy progression in Parkinson's disease using the GOrilla platform. The fold enrichment was obtained by calculating the ratio of the genes positively related to the atrophy progression after two (A) and four years (B) over the number of expected genes for each GO term, based on the background genes list. Only results with a $p\text{-}value_{Bonferonni} < 0.05$ were presented. No significant categories were found for the genes negatively related to the atrophy progression after two and four years. The GO terms from the GOrilla platform positively associated with atrophy progression after two (*N*=26) and four years (*N*=23) respectively overlap with 88% and 74% of the terms from PANTHER.

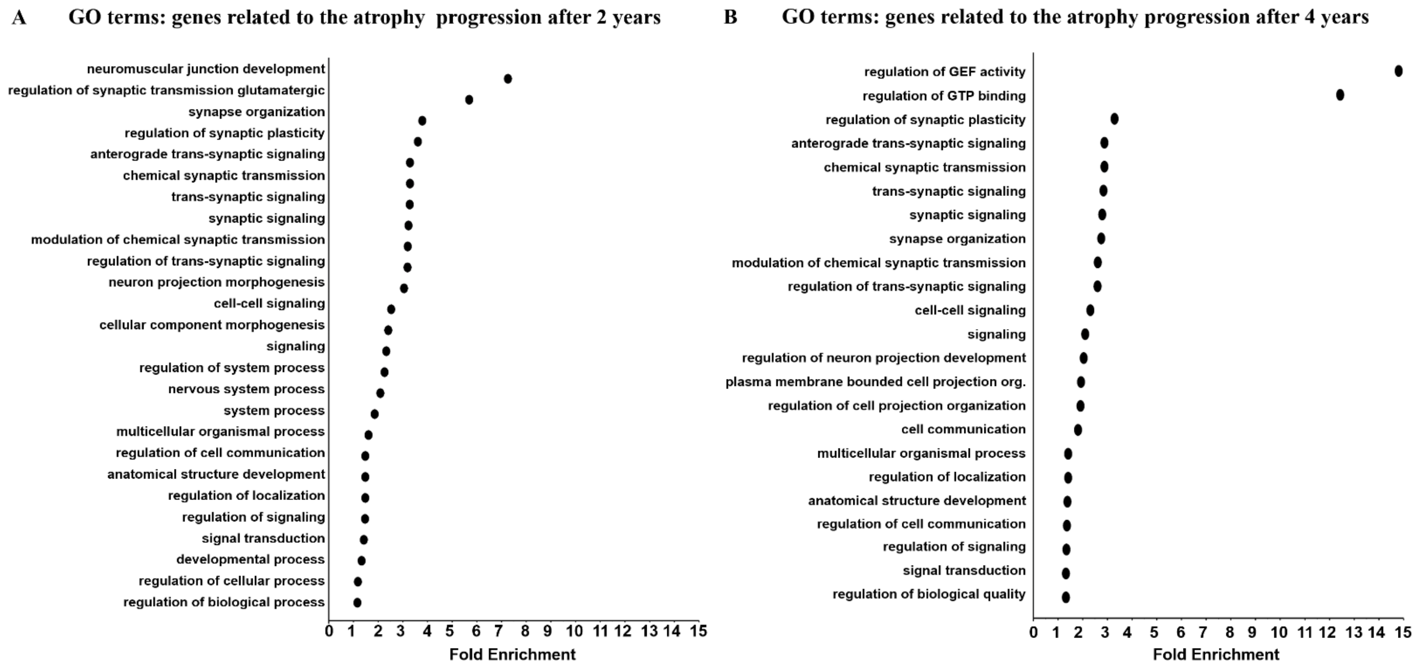

**Figure 6.** Gene ontology (GO) enrichment analysis of all the genes related to atrophy progression in Parkinson's disease using the PANTHER platform. The fold enrichment was obtained by calculating the ratio of the genes positively related to the atrophy progression after two (A) and four years (B) over the number of expected genes for each GO term, based on the background genes list. Only results with a $p\text{-}value_{Bonferonni} < 0.05$ were presented. No significant categories were found for the genes negatively related to the atrophy progression after two and four years.

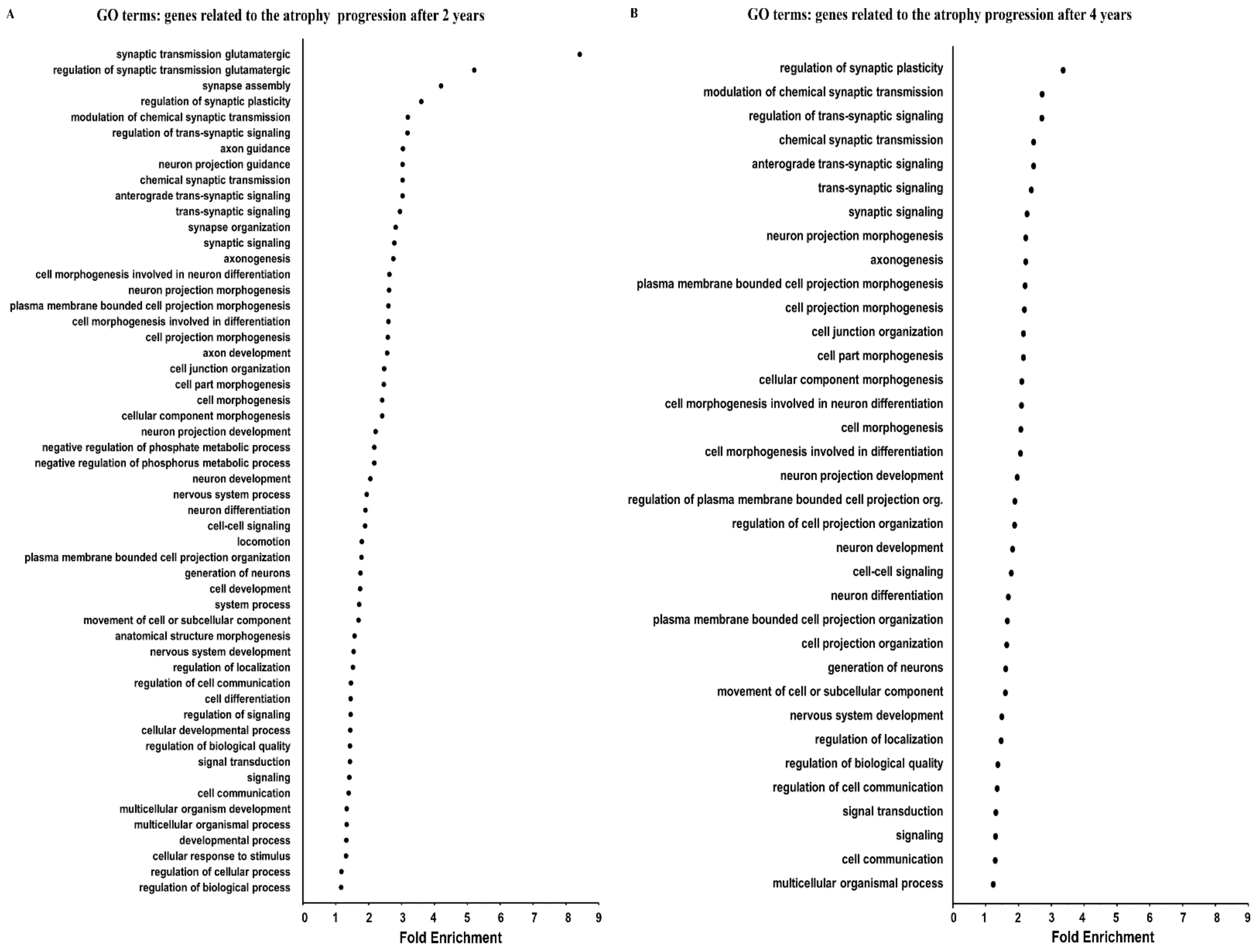

**Table 4.** Genes located in PD GWAS loci associated with each GO term related to atrophy progression after two years in *de novo* Parkinson's disease

| GO terms | Genes located in PD GWAS loci | Nearest Genes |
|---|---|---|
| Regulation of synaptic transmission glutamatergic | None | |
| Synapse organization | BSN - bassoon presynaptic cytomatrix protein | IP6K2 |
| Regulation of synaptic plasticity | PRRT2 - proline-rich transmembrane protein 2 | CD19 |
| Anterograde trans-synaptic signaling | BSN - bassoon presynaptic cytomatrix protein | IP6K2 |
| | EGR3 - early growth response 3 | BIN3 |
| Chemical synaptic transmission | BSN - bassoon presynaptic cytomatrix protein | IP6K2 |
| | EGR3 - early growth response 3 | BIN3 |
| Trans-synaptic signaling | BSN - bassoon presynaptic cytomatrix protein | IP6K2 |
| | EGR3 - early growth response 3 | BIN3 |
| Synaptic signaling | BSN - bassoon presynaptic cytomatrix protein | IP6K2 |
| | EGR3 - early growth response 3 | BIN3 |
| Modulation of chemical synaptic transmission | PRRT2 - proline-rich transmembrane protein 2 | CD19 |
| Regulation of trans-synaptic signaling | None | |
| Neuron projection morphogenesis | None | |
| Cell-cell signaling | FGFR3 - fibroblast growth factor receptor 3 | TMEM175, GAK |
| | EGR3 - early growth response 3 | BIN3 |
| Cellular component morphogenesis | SHROOM3 - shroom family member 3 | FAM47E, SCARB2, FAM47E-STBD1 |
| Signaling | BSN - bassoon presynaptic cytomatrix protein | IP6K2 |
| | EGR3 - early growth response 3 | BIN3 |
| | FGFR3 - fibroblast growth factor receptor 3 | TMEM175, GAK |
| | CAMK2D - calcium/calmodulin-dependent protein kinase ii delta | CAMK2D |
| Nervous system process | TTC8 - tetratricopeptide repeat domain 8 | GALC |
| System process | TTC8 - tetratricopeptide repeat domain 8 | GALC |
| | CAMK2D - calcium/calmodulin-dependent protein kinase ii delta | CAMK2D |

| | | |
|---|---|---|
| Multicellular organismal process | ABCA1 - atp-binding cassette sub-family a (abc1) member 1 | SLC44A1 |
| | TRIM8 - tripartite motif containing 8 | GBF1 |
| | FGFR3 - fibroblast growth factor receptor 3 | TMEM175, GAK |
| | TTC8 - tetratricopeptide repeat domain 8 | GALC |
| | CAMK2D - calcium/calmodulin-dependent protein kinase ii delta | CAMK2D |
| | SHROOM3 - shroom family member 3 | FAM47E, SCARB2, FAM47E-STBD1 |
| | SMAD4 - smad family member 4 | MEX3C |
| | TMEM107 - transmembrane protein 107 | CHRNB1 |
| Regulation of cell communication | FAM13A - family with sequence similarity 13 member a | SNCA |
| | ABCA1 - atp-binding cassette sub-family a (abc1) member 1 | SLC44A1 |
| | LGALS3 - lectin galactoside-binding soluble 3 | GCH1 |
| | UBAP2 - ubiquitin associated protein 2 | UBAP2 |
| | TRIM8 - tripartite motif containing 8 | GBF1 |
| | FGFR3 - fibroblast growth factor receptor 3 | TMEM175, GAK |
| | PDCD4 - programmed cell death 4 (neoplastic transformation inhibitor) | ADRA2A, SLC44A1 |
| | CAMK2D - calcium/calmodulin-dependent protein kinase ii delta | CAMK2D |
| | GRK5 - g protein-coupled receptor kinase 5 | INPP5F, BAG3 |
| | GPAM - glycerol-3-phosphate acyltransferase mitochondrial | ADRA2A |
| | SMAD4 - smad family member 4 | MEX3C |
| Regulation of localization | ABCA1 - atp-binding cassette sub-family a (abc1) member 1 | SLC44A1 |
| | LGALS3 - lectin galactoside-binding soluble 3 | GCH1 |
| | TRIM8 - tripartite motif containing 8 | GBF1 |
| | C5orf30 - chromosome 5 open reading frame 30 | PAM |
| | BSN - bassoon presynaptic cytomatrix protein | IP6K2 |
| | CAMK2D - calcium/calmodulin-dependent protein kinase ii delta | CAMK2D |
| | SEMA4A - sema domain immunoglobulin domain (ig) transmembrane domain (tm) and short cytoplasmic domain (semaphorin) 4a | PMVK, KRTCAP2, GBAP1 |
| | IPO5 - importin 5 | MBNL2 |
| | SMAD4 - smad family member 4 | MEX3C |
| Regulation of signaling | FAM13A - family with sequence similarity 13 member a | SNCA |
| | ABCA1 - atp-binding cassette sub-family a (abc1) member 1 | SLC44A1 |

| | | |
|---|---|---|
| | LGALS3 - lectin galactoside-binding soluble 3 | GCH1 |
| | UBAP2 - ubiquitin associated protein 2 | UBAP2 |
| | TRIM8 - tripartite motif containing 8 | GBF1 |
| | FGFR3 - fibroblast growth factor receptor 3 | TMEM175, GAK |
| | PDCD4 - programmed cell death 4 (neoplastic transformation inhibitor) | ADRA2A, SLC44A1 |
| | CAMK2D - calcium/calmodulin-dependent protein kinase ii delta | CAMK2D |
| | GRK5 - g protein-coupled receptor kinase 5 | INPP5F, BAG3 |
| | GPAM - glycerol-3-phosphate acyltransferase mitochondrial | ADRA2A |
| | SMAD4 - smad family member 4 | MEX3C |
| Signal transduction | FAM13A - family with sequence similarity 13 member a | SNCA |
| | ABCA1 - atp-binding cassette sub-family a (abc1) member 1 | SLC44A1 |
| | TRIM8 - tripartite motif containing 8 | GBF1 |
| | SERP1 - stress-associated endoplasmic reticulum protein 1 | MED12L |
| | FGFR3 - fibroblast growth factor receptor 3 | TMEM175, GAK |
| | BSN - bassoon presynaptic cytomatrix protein | IP6K2 |
| | PDCD4 - programmed cell death 4 (neoplastic transformation inhibitor) | ADRA2A, SLC44A1 |
| | CAMK2D - calcium/calmodulin-dependent protein kinase ii delta | CAMK2D |
| | SEMA4A - sema domain immunoglobulin domain (ig) transmembrane domain (tm) and short cytoplasmic domain (semaphorin) 4a | PMVK, KRTCAP2, GBAP1 |
| | PITPNM2 - phosphatidylinositol transfer protein membrane-associated 2 | HIP1R |
| | GRK5 - g protein-coupled receptor kinase 5 | INPP5F, BAG3 |
| | SMAD4 - smad family member 4 | MEX3C |
| Developmental process | LGALS3 - lectin galactoside-binding soluble 3 | GCH1 |
| | EGR3 - early growth response 3 | BIN3 |
| | TRIM8 - tripartite motif containing 8 | GBF1 |
| | PALLD - palladin cytoskeletal associated protein | CLCN3 |
| | TMEM107 - transmembrane protein 107 | CHRNB1 |
| | SMC6 - structural maintenance of chromosomes 6 | KCNS3 |
| | ASXL3 - additional sex combs like 3 (drosophila) | ASXL3 |
| | FGFR3 - fibroblast growth factor receptor 3 | TMEM175, GAK |
| | PDCD4 - programmed cell death 4 (neoplastic transformation inhibitor) | ADRA2A, SLC44A1 |

| | | |
|---|---|---|
| | SEMA4A - sema domain immunoglobulin domain (ig) transmembrane domain (tm) and short cytoplasmic domain (semaphorin) 4a | PMVK, KRTCAP2, GBAP1 |
| | SHROOM3 - shroom family member 3 | FAM47E, SCARB2, FAM47E-STBD1 |
| | SMAD4 - smad family member 4 | MEX3C |
| Regulation of cellular process | FAM13A - family with sequence similarity 13 member a | SNCA |
| | ABCA1 - atp-binding cassette sub-family a (abc1) member 1 | SLC44A1 |
| | LGALS3 - lectin galactoside-binding soluble 3 | GCH1 |
| | EGR3 - early growth response 3 | BIN3 |
| | TRIM8 - tripartite motif containing 8 | GBF1 |
| | BSN - bassoon presynaptic cytomatrix protein | IP6K2 |
| | ZNF681 - zinc finger protein 681 | TMEM175, GAK |
| | CAMK2D - calcium/calmodulin-dependent protein kinase ii delta | CAMK2D |
| | IPO5 - importin 5 | MBNL2 |
| | CELF5 - cugbp elav-like family member 5 | SPPL2B |
| | CCNG2 - cyclin g2 | FAM47E, SCARB2, FAM47E-STBD1 |
| | UBTF - upstream binding transcription factor rna polymerase i | UBTF, FAM171A2 |
| | YEATS2 - yeats domain containing 2 | MCCC1 |
| | SMC6 - structural maintenance of chromosomes 6 | KCNS3 |
| | ASXL3 - additional sex combs like 3 (drosophila) | ASXL3 |
| | ZNF184 - zinc finger protein 184 | LOC100131289 |
| | UBAP2 - ubiquitin associated protein 2 | UBAP2 |
| | SERP1 - stress-associated endoplasmic reticulum protein 1 | MED12L |
| | FGFR3 - fibroblast growth factor receptor 3 | TMEM175, GAK |
| | C5orf30 - chromosome 5 open reading frame 30 | PAM |
| | PDCD4 - programmed cell death 4 (neoplastic transformation inhibitor) | ADRA2A, SLC44A1 |
| | SEMA4A - sema domain immunoglobulin domain (ig) transmembrane domain (tm) and short cytoplasmic domain (semaphorin) 4a | PMVK, KRTCAP2, GBAP1 |
| | PITPNM2 - phosphatidylinositol transfer protein membrane-associated 2 | HIP1R |
| | GRK5 - g protein-coupled receptor kinase 5 | INPP5F, BAG3 |
| | SHROOM3 - shroom family member 3 | FAM47E, SCARB2, FAM47E-STBD1 |
| | ZNF26 - zinc finger protein 26 | FBRSL1 |

| | | |
|---|---|---|
| | GPAM - glycerol-3-phosphate acyltransferase mitochondrial | ADRA2A |
| | SMAD4 - smad family member 4 | MEX3C |
| Regulation of biological process | ABCA1 - atp-binding cassette sub-family a (abc1) member 1 | SLC44A1 |
| | LGALS3 - lectin galactoside-binding soluble 3 | GCH1 |
| | IPO5 - importin 5 | MBNL2 |
| | CELF5 - cugbp elav-like family member 5 | SPPL2B |
| | YEATS2 - yeats domain containing 2 | MCCC1 |
| | SMC6 - structural maintenance of chromosomes 6 | KCNS3 |
| | ZNF184 - zinc finger protein 184 | LOC100131289 |
| | C5orf30 - chromosome 5 open reading frame 30 | PAM |
| | PITPNM2 - phosphatidylinositol transfer protein membrane-associated 2 | HIP1R |
| | GRK5 - g protein-coupled receptor kinase 5 | INPP5F, BAG3 |
| | SHROOM3 - shroom family member 3 | FAM47E, SCARB2, FAM47E-STBD1 |
| | ZNF26 - zinc finger protein 26 | FBRSL1 |
| | GPAM - glycerol-3-phosphate acyltransferase mitochondrial | ADRA2A |
| | SMAD4 - smad family member 4 | MEX3C |
| | FAM13A - family with sequence similarity 13 member a | SNCA |
| | EGR3 - early growth response 3 | BIN3 |
| | TRIM8 - tripartite motif containing 8 | GBF1 |
| | BSN - bassoon presynaptic cytomatrix protein | IP6K2 |
| | ZNF876P - zinc finger protein 876 pseudogene | TMEM175, GAK |
| | CAMK2D - calcium/calmodulin-dependent protein kinase ii delta | CAMK2D |
| | CCNG2 - cyclin g2 | FAM47E, SCARB2, FAM47E-STBD1 |
| | UBTF - upstream binding transcription factor rna polymerase i | UBTF, FAM171A2 |
| | ASXL3 - additional sex combs like 3 (drosophila) | ASXL3 |
| | UBAP2 - ubiquitin associated protein 2 | UBAP2 |
| | SERP1 - stress-associated endoplasmic reticulum protein 1 | MED12L |
| | FGFR3 - fibroblast growth factor receptor 3 | TMEM175, GAK |
| | PDCD4 - programmed cell death 4 (neoplastic transformation inhibitor) | ADRA2A, SLC44A1 |
| | SEMA4A - sema domain immunoglobulin domain (ig) transmembrane domain (tm) and short cytoplasmic domain (semaphorin) 4a | PMVK, KRTCAP2, GBAP1 |

**Table 5.** Genes located in PD GWAS loci associated with each GO term related to atrophy progression after four years in *de novo* Parkinson's disease

| GO terms | Genes located in PD GWAS loci | Nearest Genes |
|---|---|---|
| Regulation of synaptic plasticity | RIMS1 - regulating synaptic membrane exocytosis 1 | RIMS1 |
| | MAPT - microtubule-associated protein tau | CRHR1, WNT3 |
| Anterograde trans-synaptic signaling | BSN - bassoon presynaptic cytomatrix protein | IP6K2 |
| | EGR3 - early growth response 3 | BIN3 |
| | P2RX2 - purinergic receptor p2x ligand-gated ion channel 2 | FBRSL1 |
| | DOC2A - double c2-like domains alpha | SETD1A |
| Chemical synaptic transmission | BSN - bassoon presynaptic cytomatrix protein | IP6K2 |
| | DOC2A - double c2-like domains alpha | SETD1A |
| | EGR3 - early growth response 3 | BIN3 |
| | P2RX2 - purinergic receptor p2x ligand-gated ion channel 2 | FBRSL1 |
| Trans-synaptic signaling | BSN - bassoon presynaptic cytomatrix protein | IP6K2 |
| | DOC2A - double c2-like domains alpha | SETD1A |
| | EGR3 - early growth response 3 | BIN3 |
| | P2RX2 - purinergic receptor p2x ligand-gated ion channel 2 | FBRSL1 |
| Synaptic signaling | BSN - bassoon presynaptic cytomatrix protein | IP6K2 |
| | DOC2A - double c2-like domains alpha | SETD1A |
| | EGR3 - early growth response 3 | BIN3 |
| | P2RX2 - purinergic receptor p2x ligand-gated ion channel 2 | FBRSL1 |
| Modulation of chemical synaptic transmission | MAPT - microtubule-associated protein tau | CRHR1, WNT3 |
| | RIMS1 - regulating synaptic membrane exocytosis 1 | RIMS1 |
| Regulation of trans-synaptic signaling | MAPT - microtubule-associated protein tau | CRHR1, WNT3 |
| | RIMS1 - regulating synaptic membrane exocytosis 1 | RIMS1 |
| Cell-cell signaling | BSN - bassoon presynaptic cytomatrix protein | IP6K2 |
| | DOC2A - double c2-like domains alpha | SETD1A |
| | EGR3 - early growth response 3 | BIN3 |
| | FGFR3 - fibroblast growth factor receptor 3 | TMEM175, GAK |
| | MAPT - microtubule-associated protein tau | CRHR1, WNT3 |

| | | |
|---|---|---|
| | P2RX2 - purinergic receptor p2x ligand-gated ion channel 2 | FBRSL1 |
| Signaling | BSN - bassoon presynaptic cytomatrix protein | IP6K2 |
| | DOC2A - double c2-like domains alpha | SETD1A |
| | EGR3 - early growth response 3 | BIN3 |
| | FGFR3 - fibroblast growth factor receptor 3 | TMEM175, GAK |
| | CAMK2D - calcium/calmodulin-dependent protein kinase ii delta | CAMK2D |
| | MAPT - microtubule-associated protein tau | CRHR1, WNT3 |
| | P2RX2 - purinergic receptor p2x ligand-gated ion channel 2 | FBRSL1 |
| Regulation of plasma membrane bounded cell projection organization | SEMA4A - sema domain immunoglobulin domain (ig) transmembrane domain (tm) and short cytoplasmic domain (semaphorin) 4a | PMCK, KRTCAP2, GBAP1 |
| | RIMS1 - regulating synaptic membrane exocytosis 1 | RIMS1 |
| | MAPT - microtubule-associated protein tau | CRHR1, WNT3 |
| | TAPT1 - transmembrane anterior posterior transformation 1 | BST1 |
| Regulation of cell projection organization | MAPT - microtubule-associated protein tau | CRHR1, WNT3 |
| | RIMS1 - regulating synaptic membrane exocytosis 1 | RIMS1 |
| | SEMA4A - sema domain immunoglobulin domain (ig) transmembrane domain (tm) and short cytoplasmic domain (semaphorin) 4a | PMCK, KRTCAP2, GBAP1 |
| | TAPT1 - transmembrane anterior posterior transformation 1 | BST1 |
| Cell communication | BSN - bassoon presynaptic cytomatrix protein | IP6K2 |
| | DOC2A - double c2-like domains alpha | SETD1A |
| | EGR3 - early growth response 3 | BIN3 |
| | FGFR3 - fibroblast growth factor receptor 3 | TMEM175, GAK |
| | MAPT - microtubule-associated protein tau | CRHR1, WNT3 |
| | P2RX2 - purinergic receptor p2x ligand-gated ion channel 2 | FBRSL1 |
| Multicellular organismal process | CAMK2D - calcium/calmodulin-dependent protein kinase ii delta | CAMK2D |
| | ABCA1 - atp-binding cassette sub-family a (abc1) member 1 | SLC44A1 |
| | FGFR3 - fibroblast growth factor receptor 3 | TMEM175, GAK |
| | BTRC - beta-transducin repeat containing e3 ubiquitin protein ligase | GBF1 |
| | MAPT - microtubule-associated protein tau | CRHR1, WNT3 |
| | LEFTY1 - left-right determination factor 1 | ITPKB |
| | NMT1 - n-myristoyltransferase 1 | UBTF, FAM171A2, CRHR1 |

| | | |
|---|---|---|
| | P2RX2 - purinergic receptor p2x ligand-gated ion channel 2 | FBRSL1 |
| | RIMS1 - regulating synaptic membrane exocytosis 1 | RIMS1 |
| | SHROOM3 - shroom family member 3 | FAM47E, SCARB2, FAM47E-STBD1 |
| | RUNDC1 - run domain containing 1 | RETREG3 |
| | SMAD4 - smad family member 4 | MEX3C |
| | TRIM8 - tripartite motif containing 8 | GBF1 |
| | TTC8 - tetratricopeptide repeat domain 8 | GALC |
| | UBE2B - ubiquitin-conjugating enzyme e2b | C5orf24 |
| | TAPT1 - transmembrane anterior posterior transformation 1 | BST1 |
| | TMEM107 - transmembrane protein 107 | CHRNB1 |
| Regulation of localization | ABCA1 - atp-binding cassette sub-family a (abc1) member 1 | SLC44A1 |
| | BSN - bassoon presynaptic cytomatrix protein | IP6K2 |
| | CAMK2D - calcium/calmodulin-dependent protein kinase ii delta | CAMK2D |
| | DOC2A - double c2-like domains alpha | SETD1A |
| | MAPT - microtubule-associated protein tau | CRHR1, WNT3 |
| | NMT1 - n-myristoyltransferase 1 | UBTF, FAM171A2, CRHR1 |
| | LGALS3 - lectin galactoside-binding soluble 3 | GCH1 |
| | IPO5 - importin 5 | MBNL2 |
| | P2RX2 - purinergic receptor p2x ligand-gated ion channel 2 | FBRSL1 |
| | RIMS1 - regulating synaptic membrane exocytosis 1 | RIMS1 |
| | SEMA4A - sema domain immunoglobulin domain (ig) transmembrane domain (tm) and short cytoplasmic domain (semaphorin) 4a | PMCK, KRTCAP2, GBAP1 |
| | SMAD4 - smad family member 4 | MEX3C |
| | TRIM8 - tripartite motif containing 8 | GBF1 |
| | STX7 - syntaxin 7 | RPS12 |
| Regulation of cell communication | ABCA1 - atp-binding cassette sub-family a (abc1) member 1 | SLC44A1 |
| | CAMK2D - calcium/calmodulin-dependent protein kinase ii delta | CAMK2D |
| | BTRC - beta-transducin repeat containing e3 ubiquitin protein ligase | GBF1 |
| | BRD7 - bromodomain containing 7 | NOD2 |
| | FGFR3 - fibroblast growth factor receptor 3 | TMEM175, GAK |
| | FAM13A - family with sequence similarity 13 member a | SNCA |

| | | |
|---|---|---|
| | GPAM - glycerol-3-phosphate acyltransferase mitochondrial | ADRA2A |
| | MAPT - microtubule-associated protein tau | CRHR1, WNT3 |
| | LGALS3 - lectin galactoside-binding soluble 3 | GCH1 |
| | LEFTY1 - left-right determination factor 1 | ITPKB |
| | GRK5 - g protein-coupled receptor kinase 5 | INPP5F, BAG3 |
| | P2RX2 - purinergic receptor p2x ligand-gated ion channel 2 | FBRSL1 |
| | NMT1 - n-myristoyltransferase 1 | UBTF, FAM171A2, CRHR1 |
| | PYDC1 - pyd (pyrin domain) containing 1 | SETD1A |
| | RIMS1 - regulating synaptic membrane exocytosis 1 | RIMS1 |
| | SMAD4 - smad family member 4 | MEX3C |
| | TRIM8 - tripartite motif containing 8 | GBF1 |
| | SH3RF1 - sh3 domain containing ring finger 1 | CLCN3 |
| | UBE2B - ubiquitin-conjugating enzyme e2b | C5orf24 |
| | UBAP2 - ubiquitin associated protein 2 | UBAP2 |
